# Facilitating clinically relevant skin tumor diagnostics with spectroscopy-driven machine learning

**DOI:** 10.1101/2023.10.14.23296584

**Authors:** Emil Andersson, Jenny Hult, Carl Troein, Magne Stridh, Benjamin Sjögren, Agnes Pekar-Lukacs, Julio Hernandez-Palacios, Patrik Edén, Bertil Persson, Victor Olariu, Malin Malmsjö, Aboma Merdasa

## Abstract

In the dawning era of artificial intelligence (AI), health care stands to undergo a significant transformation with the increasing digitalization of patient data. Digital imaging, in particular, will serve as an important platform for AI to be implemented to aid decision making and diagnostics. A growing number of studies demonstrate the potential of AI for automatic pre-surgical skin tumor delineation, which could have tremendous impact on clinical practice. However, current methods have the drawback of relying on a ground truth image in which the tumor borders are already identified, which is not clinically possible. We report a novel approach where hyperspectral images provides spectra from small regions representing healthy tissue and tumor, which are used to generate prediction maps using artificial neural networks. Thereafter, a segmentation algorithm automatically manages to determine the skin tumor borders. Our approach therefore circumvents the need for a complete ground truth image, where the training data is contained within each individual patient. This links to an important strength of our approach as we develop individual network models for each patient. Our approach is therefore not only more clinically relevant, but it also interesting for emerging precision skin tumor diagnostics where adaptability toward the individual is key.

## Introduction

With over 1.5 million new cases in 2020, skin cancer is the third most common cancer worldwide.^1^ Non-melanoma skin cancer (NMSC) is the most common group of cancers in western countries,^2^ comprising basal cell carcinoma (BCC) and squamous cell carcinomas (SCC). Malignant melanoma is more rare than NMSC, but is also more aggressive and represents a growing disease burden in society.^3,4^ Current clinical practice to diagnose and treat skin tumors is unfortunately rather time-consuming, involving excisional biopsy and histopathological analysis. With an increasing incidence worldwide,^1^ there is a justified concern that a seemingly cumbersome diagnostic procedure may cause a bottleneck for the growing diagnostic demand.

There have been several reports over recent years implementing artificial intelligence (AI) and machine learning (ML) methods with dermatoscopy to facilitate more efficient diagnosis.^5,6^ These methods offer pre-surgical skin tumor diagnostics which have high clinical impact AI-based applications have emerged for mobile and hand-held devices to assess malignancy of lesions and guide decision making in whether to perform a biopsy or apply other treatment measures.^7,8^ However, while there is an obvious benefit of early classification of a suspected lesion, in order to improve survival rate, identifying skin tumor borders is often overlooked although it is of high clinical importance relating to treatment and prognosis. Despite guidelines to ensure complete skin tumor removal,^9,10^ up toward 78% of primary biopsies are discovered via histopathology to be non-radical,^11^ which consequently requires re-surgery incurring additional health care costs and patient suffering, while unfortunately also reducing survivability.^12^

Driven by the clinical need for reducing the number of non-radical biopsy excisions, studies employing artificial neural networks (ANNs) to pre-surgically delineate skin tumors in order to assist surgeons are emerging.^13^ However, they suffer a few limitations considering their clinical applicability. Performance of AI models employing feature extraction depend heavily on image resolution.^14^ This may also be affected by patient motion during image acquisition. A recent report also demonstrates the relative ease at which a convolutional neural network (CNN) image classifier produces inconsistent predictions when simply rotating a dermascopic tumor image.^15^ These examples underline the potential pitfalls of image classifiers that rely on spatial feature extraction.

Another major limitation for AI and ML implementations for skin tumor delineation is tied to how image classifiers fundamentally operate. Previous reports employing an image classifier to automatically identify skin tumor borders conventionally require training images where the tumor borders have been manually identified.^16,17^ From a clinical perspective, this poses a challenge since the only reliable way to currently determine the border between healthy tissue and skin tumor is via histopathology. By chemically staining biopsy cross-sections, the spectral (color) contrast is enhanced such that healthy tissue and tumor can easily be differentiated, which otherwise would not be observable via direct visual inspection or using a dermatoscope.^18,19^ While AI models certainly can automate the skin tumor delineation to a level of accuracy comparable to a clinician manually drawing them in dermoscopic images,^20–22^ the real question is whether this is sufficient for clinical implementation since histopathology still is required. If an image does not contain the necessary spectral contrast in order to produce a clinically reliable diagnosis,^23^ the data quality is automatically insufficient and alternative approaches should be considered.^24,25^

In this work we present an alternative ML approach to pre-surgical skin tumor delineation that does not require manual identification of the tumor borders. Rather than analyzing spatial features in standard color images, our model instead builds on recognizing spectral patterns in hyperspectral images^26–28^ containing information beyond the sensitivity of the naked eye. We detail a model that trains on the spectra from image regions that belong to healthy tissue or skin tumor, which enables us to automatically determine and visualize the border between healthy tissue and tumor. A key advantage of the presented model is that the training data is contained within each individual patient, which has important clinical implications. Moreover, exploiting the ability to generate individualized ANN models for each sample, we demonstrate the adaptability of the approach to use the extended spectral contrast between healthy tissue and tumor for different patients and skin tumors types. This taps into the potential of using the presented approach for personalized medicine.

## Results & Discussion

### Different spectra for healthy and tumor pixels

Figure 1 shows a representative example of the information that can be extracted from a hyperspectral image (see Methods for instrument details). A regular color photo of a melanoma tumor sample is used as a reference (Figure 1a). Absorbance spectra are extracted from every pixel (averaged vertically over +/- 5 pixels) along the horizontal white dotted line spanning from and to healthy tissue across the tumor. Figure 1b shows all these spectra combined into a heat map where it is evident there is spectral contrast comparing regions representing healthy tissue and tumor. This is most evident in the 600-1000 nm spectral range. However, some contrast can also be observed for longer wavelengths. Figure 1c shows three individual spectra extracted from the locations indicated in Figures 1a-b, where the spectral contrast becomes more evident. Figure 1d shows three examples of false color images that can be produced from the hyperspectral image data set using a combination of monochromatic images from three unique spectral channels. From the 322 spectral channels available in the data set, over 33 million unique false color images can in essence be created, which demonstrates the flexibility in image representation offered by our system. The three examples in Figure 1d highlight how the hyperspectral information can be used to display potentially clinically relevant information. The top image combines three spectral channels that represent the background information without any sign of the tumor. The middle image shows the tumor clearly, while the bottom image shows the tumor together with two additional spots presumed to be blood clots. While a trained clinician likely can differentiate between these different regions with the naked eye, the ability of HSI to completely separate and display these regions is not only important for conventional machine learning approaches based on spatial contrasts,^29^ but also for the spectroscopically-guided machine learning methods discussed in this work.

**Figure 1.**
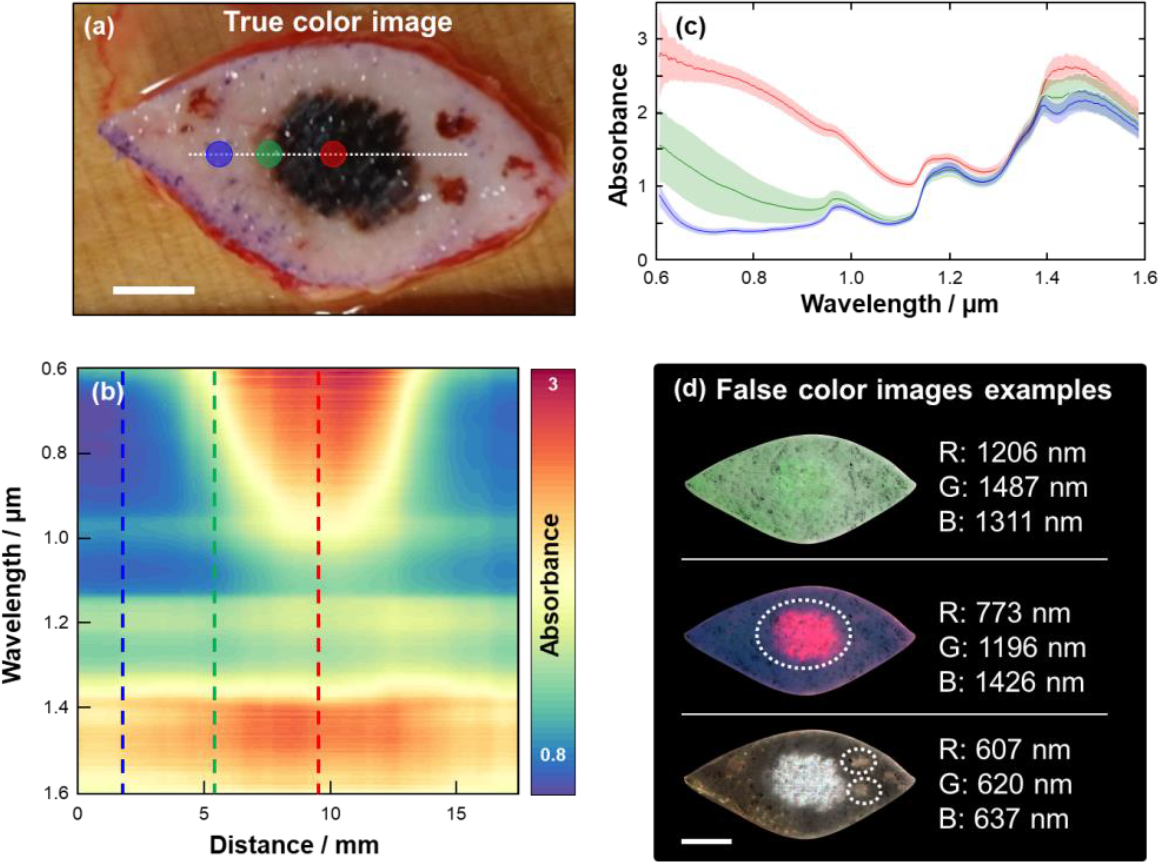
(a) Representative example of a color photo of a melanoma tumor. (b) Spectra extracted from every point along the dashed line in (a) with 50 um resolution plotted as a heat map. (c) Spectra extracted at three locations indicated in both (a) and (b) where blue likely represents healthy tissue, red represents the tumor and green some intermediate state in-between. (d) False color images of the tumor generated by compiling a subset of three images from the hyperspectral imaging data to enhance different sample features. In the middle false color image, the suspected tumor is indicated with a white dashed line that can be differentiated from the blood clots identified in the bottom false color image. The scale bars represent 10 mm in all images.

### Spectral analysis using MCR-ALS

Figure 1 provides insight into the abundance of spectral information that can yield a spatial contrast between tumor and healthy tissue. We employ multivariate curve resolution – alternating least squares (MCR-ALS)^30^ analysis to systematically extract the most common spectral features from all tumors combined (see Methods and SI for further details). We refrain from starting with an analysis based on the tumor type since we do not have enough of each to minimize the expected effects of patient-to-patient variation. We assume here that the difference in spectral features for all tumors compared with healthy tissue will be larger than the patient-to-patient variation of healthy tissue. Figure 2a shows the three most common spectral features from all tumors collectively. This suggests that all tumors can to some satisfying degree be described by a linear combination of these three spectral components. Figure 2b shows an example of three different tumor types (SCC, BCC and melanoma) that are shown in images that depict the contribution from each spectral component (see SI Note III for all tumors). In all examples, some contrast can be observed between healthy tissue and the regions considered to represent tumor. However, the best contrast is observed for different spectral components depending on the tumor type, which unsurprisingly suggests that the different tumors have different spectral features. As mentioned, however, we cannot at this stage characterize these differences in detail. The MCR-ALS algorithm could also identify four skin tumor measurements with insufficient signal-to-noise ratio (SNR) to reliably run the proceeding steps. These skin tumors were therefore excluded in the final comparison to histopathology, although the results from all analysis of each are found in the SI.

**Figure 2.**
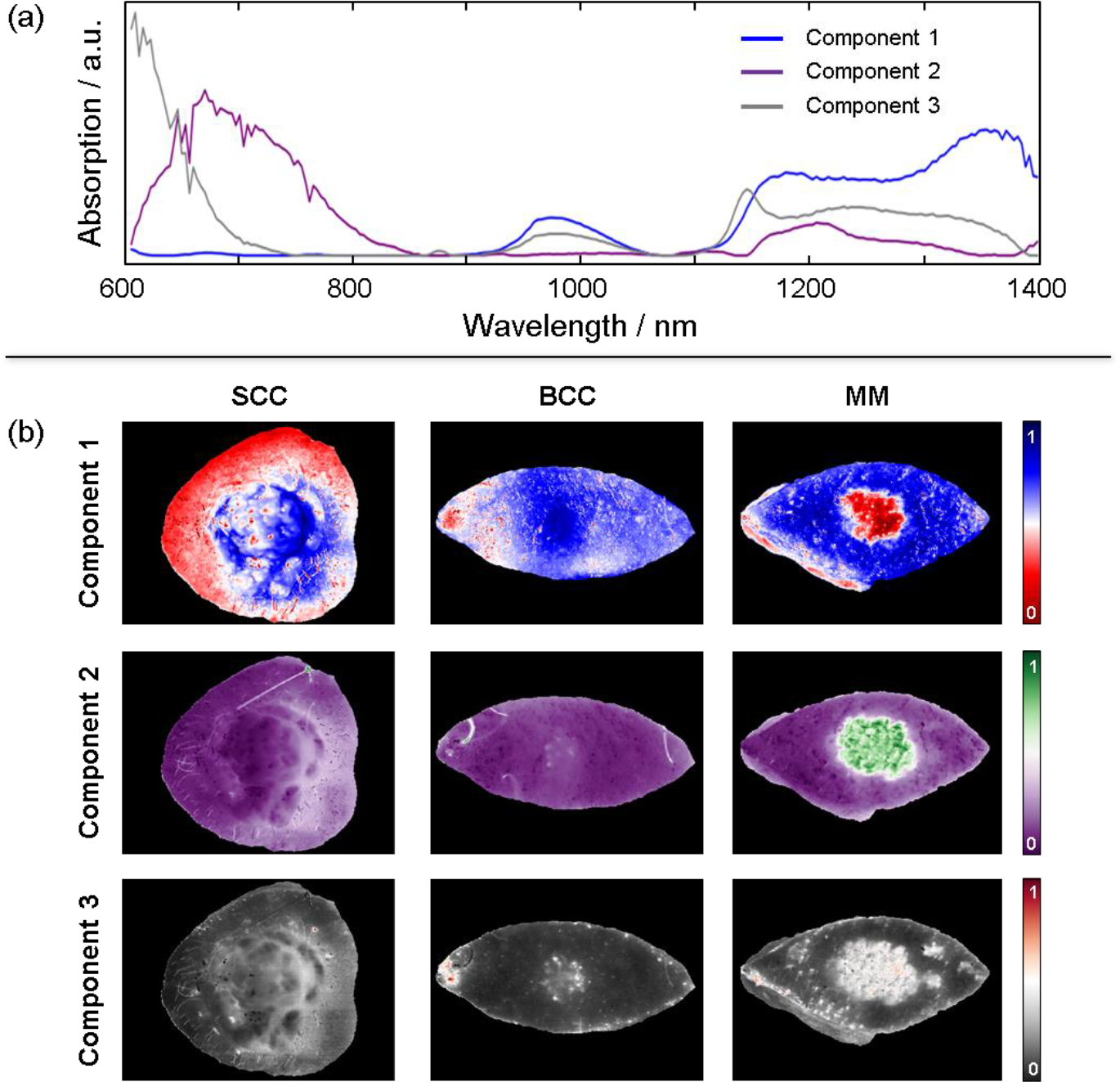
(a) Three most common spectral features (components) extracted from all tumor spectra using MCR-ALS. (b) Three examples demonstrating how new images can be produced using each spectral component depicting the relative contribution of each spectral component represented in each pixel spectrum in different colors. The three examples show how different features become visible for different spectral components depending on the tumor type.

### Spectroscopically-guided pixel prediction by machine learning

Under the assumption that there is a difference between spectra representing healthy tissue and tumor (Figure 1), an ANN-classifier should be able to learn to distinguish between the two. However, since it is not clearly known where the tumor borders are located by visual inspection of the samples, standard deep learning methods for segmentation, e.g. the U-net,^31^ are not applicable. This type of network requires annotation for every individual pixel. Hence, we chose the approach to classify each pixel individually instead of treating the image as a whole unit. Even though the exact delineation is not known, we take advantage of the fact that one can easily identify the center of the tumor visually. We select pixels from the center area of the tumor and label them as tumor pixels with high certainty. In a similar manner, pixels from edge areas of the sample, where it is clear that the tumor is not located, and there is no influence from the incision, are selected and labeled with high certainty as healthy pixels (Figure 3a). These regions were confirmed via histopathology to have been correctly categorized. The selection of tumor and healthy pixels produced two distinct training spectra in most of our samples (Figure 3b). The average spectrum for the tumor pixels in general do not overlap with the average spectrum for the selection of healthy pixels, which suggests that classification is possible based on spectral features.^32^ We note that the SNR (signal to noise ratio) in the spectral range between 1400-1600 nm is significantly lower than the rest and was therefore excluded from the spectral analysis.

**Figure 3.**
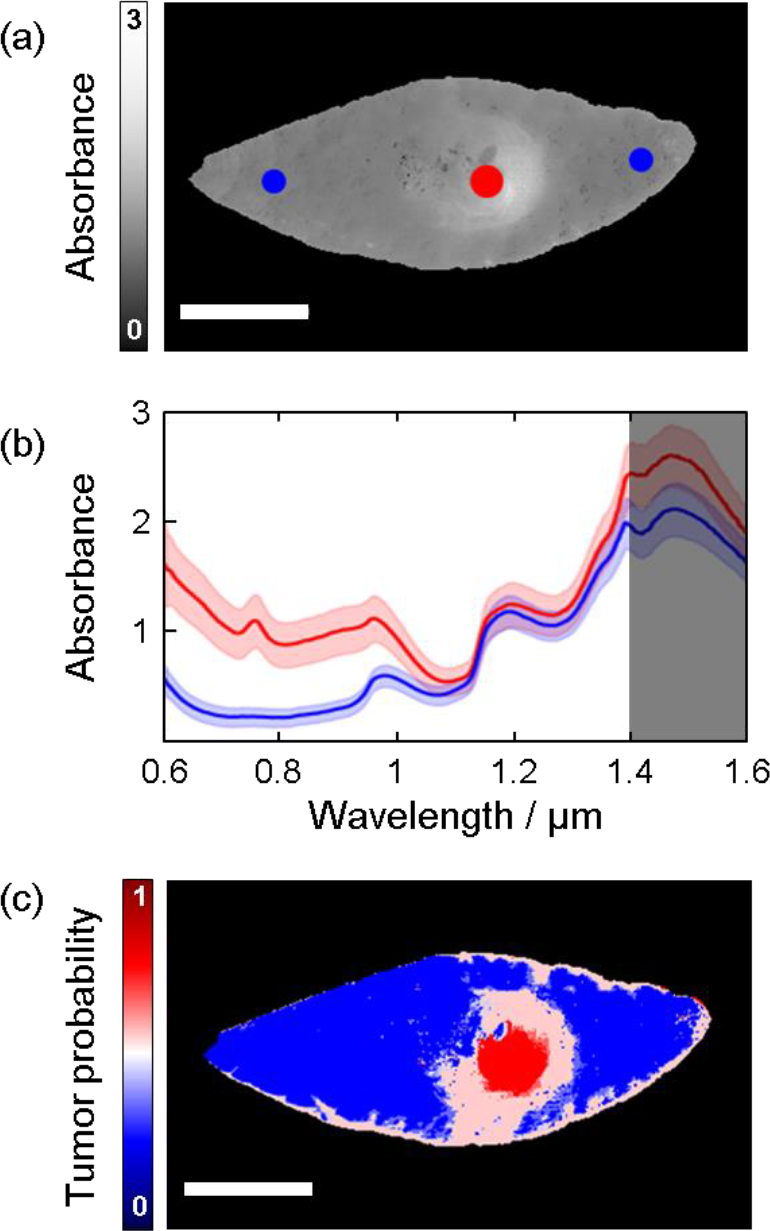
(a) Spectrally averaged image with regions representing healthy tissue (blue) and tumor (red) indicated in colored circles. (b) Average spectra extracted from the regions indicated in (a). Due to the lower SNR in the spectral range between 1400-1600 nm (dark shaded region), this range was excluded in the MCR-ALS analysis. (c) Prediction map of a malignant melanoma depicting tumor probability where red represents a 100% probability and blue represents 0% probability. The scale bars represent 10 mm.

We devised three different Neural Network models as briefly described in Methods below (see SI Note I for full description). A multilayer perceptron (MLP) and a 1-D convolutional neural network (CNN) were trained on the raw spectral features and an additional MLP was trained on the MCR-ALS decomposition of the spectra. For each of the three models, every pixel of the tumor samples was classified individually. An MLP prediction map for the tumor was constructed from the model outputs (Figure 3c) where each pixel is assigned a value between 0 (100% predicted probability being healthy tissue) and 1 (100% predicted probability being tumor). From this, a colormap is generated showing blue (red) if a pixel has a higher probability of representing healthy tissue (tumor). White represents an equal probability being classified as either healthy tissue or tumor. We refer the reader to SI Note IV for prediction maps of all tumors.

Viewing the prediction maps (SI Figure S6-S8) it is clear that they are not perfect representations of the tumors. In many cases, the prediction maps contain noise, which likely do not represent the tumor, but rather hair, marker pen used by the surgeon to identify the tumor, or a suture used to maintain the excised sample in place, which need to be removed.^33,34^ Moreover, the incision appears to yield some degree of miss-classification, which is caused by the exposure of sub-cutaneous fat and blood yielding contrasting spectral features to that of healthy tissue. Miss-classification originating from incision artifacts will be reduced by imaging the tumors *in situ* in the future. In addition, segmentation will be an important step to reduce the influence of noise and artifacts, particularly those that are in a close proximity to the tumor, when determining its border to healthy tissue.

### Segmentation

Since the spatial information contained in the data has hitherto not been utilized, we employ a segmentation algorithm to determine the tumor delineation. The individual pixels need to be put into context according to the pixels in their neighborhood. Furthermore, we must require from any segmentation algorithm which we may devise that it removes prediction noise and artifacts (e.g., blood stains and hair).

We chose an active contour algorithm since they are well suited to find borders of objects with irregular size.^35^ The general approach of an active contour segmentation algorithm is that an active contour, which we refer to as a *rope*, tracks the contour of an object in an image. This is achieved by constructing an energy function which is minimized. The energy function has contributions both from internal properties of the rope such as stretching and curvature, and external properties derived from the image such as intensity. Furthermore, this type of algorithm offers the flexibility to choose the energy function to minimize, which makes it possible to adapt it to suit our specific problem.

A complication is that the prediction map is not directly suitable for an active contour algorithm. The quantity corresponding to intensity in a prediction map is the tumor probability, which does not provide enough context to a tumor pixel’s location within the tumor. Although the tumor probability may be lower near the edge of the tumor in some cases, that is not always so. Furthermore, noise and artifacts can also obtain high tumor probability. Thus, we first need to devise a more suitable intensity function from the prediction map, which should reflect a tumor pixel’s spatial location within the tumor. The sand-pile method (see Methods) becomes ideal as it ensures that small regions of pixels misclassified as tumor (artifacts caused by e.g., blood, hair, marker pen or noise) will yield smaller sand-piles than large regions of pixels correctly classified as tumor. We refer the reader to SI Note II for more details.

The resulting sand-pile landscape (Figures 4a-b) is used by the active contour algorithm. The rope is initialized along the edge of the sample (Figure 4c) and the energy is minimized as described in Methods. The parameters of the rope have been balanced in such a way that the rope can pass over small sand-piles corresponding to artifacts, but not the tumor itself. They have also been balanced so that the rope does not track every small dent of the tumor outline to avoid unnecessary over-fitting. Supplementary video 1, from which a series of snapshots are shown in Figure 4c, demonstrates the active contour algorithm starting at the border of the sample and finally reaching an equilibrium position circumventing what it considers to be the tumor. Figure 4d show the final equilibrium position of the rope for three representative examples of different tumor types using different models together with the active contour algorithm to identify the tumor borders. We refer the reader to SI Note V for results on all tumors using the three different models.

**Figure 4.**
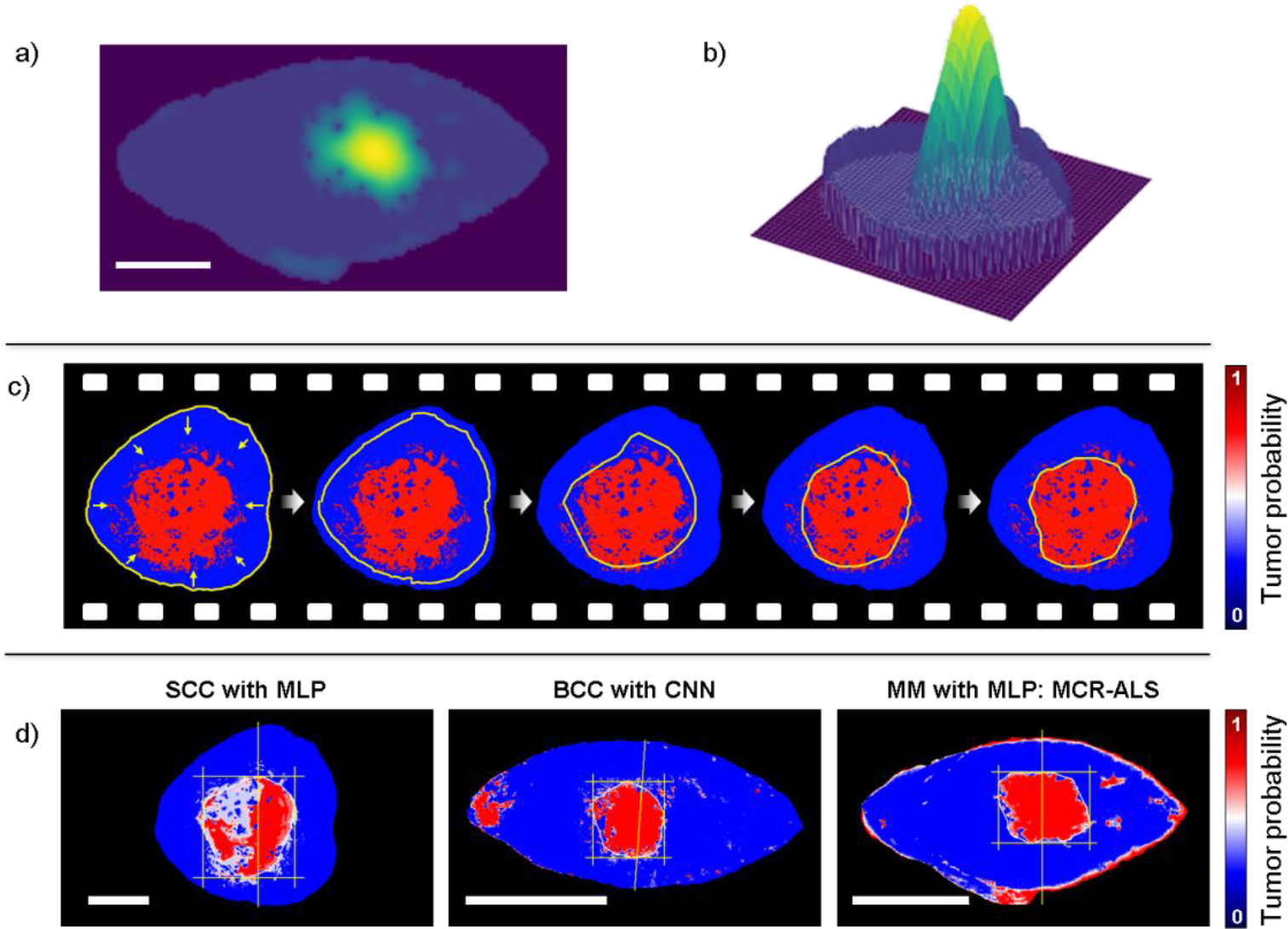
Results after performing sand-piling of a melanoma skin tumor represented as (a) a 2D image and (b) a 3D contour plot, where in the latter the difference in height becomes evident. (c) Snapshots of the active contour algorithm identifying the tumor borders by shrinking a rope from the sample borders toward the center where the tumor is. (d) Results obtained of the tumor borders of three different skin tumor types using different algorithms to generate the prediction maps. The scale bars all represent 10 mm.

### Comparison to histopathology

During histopathological analysis, we track the orientation of the cross-section cuts in relation to the sample, as well as the location at which the cross-section used to make the diagnosis is taken from. These are indicated with a vertical line as shown in Figure 4d. These are then the locations at which we extract the model predicted tumor widths in order to compare to histopathological findings. Figure 6 demonstrates the correlation between all tumor widths obtained with the three models: MLP (Figure 5a), CNN (Figure 5b) and MLP with MCR-ALS (Figure 5c). For each model, the Pearson correlation coefficient and relative mean squared error (MSE) were calculated. The relative MSE was used rather than the standard MSE since the tumors vary in size. The correlation coefficient for all three models is high, however, the tumor widths correlate differently depending on the model. This makes sense in light of the results demonstrated in Figure 3 where different prediction maps of different tumor types depict significant variations depending on which spectral component is used to contrast the spectral changes.

**Figure 5.**
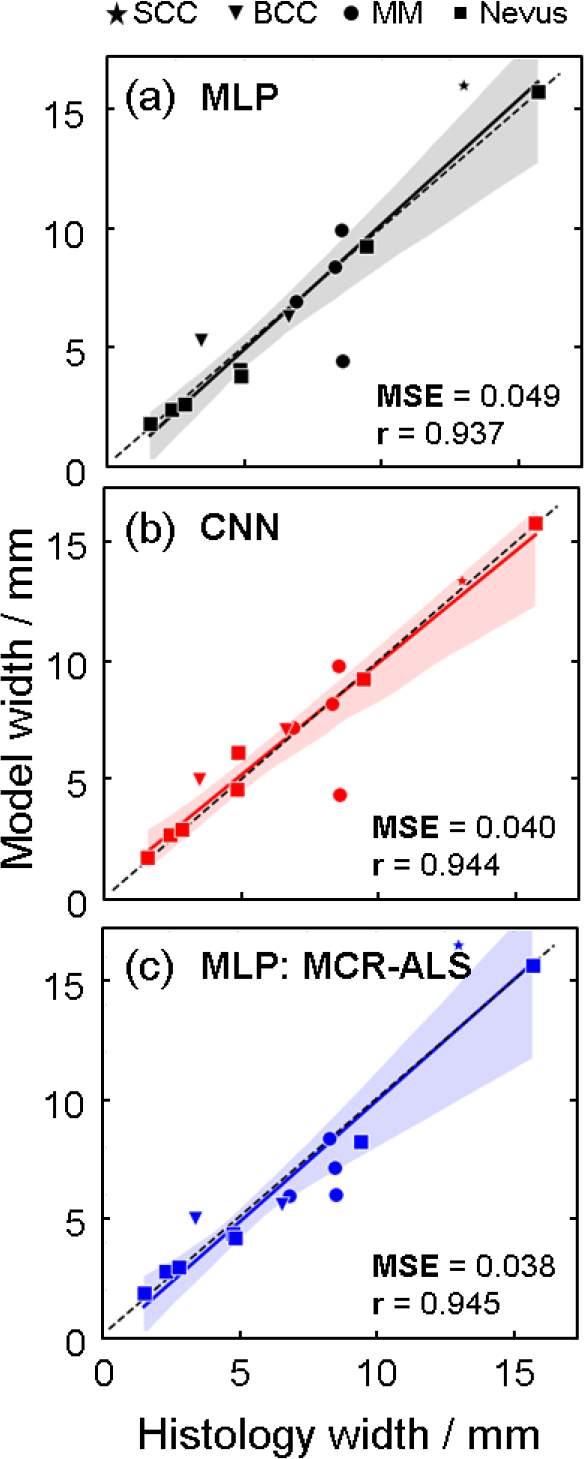
Comparison between the histopathologically determined tumor widths and those obtained by (a) MLP, (b) CNN, and (c) MLP combined with MCR-ALS. In each panel, the Pearson correlation coefficient (r) is calculated and visualized with a solid and colored trace, where the black dashed trace indicates the ground truth. The relative mean squared error (MSE) is calculated for all correlation plots.

**Figure 6.**
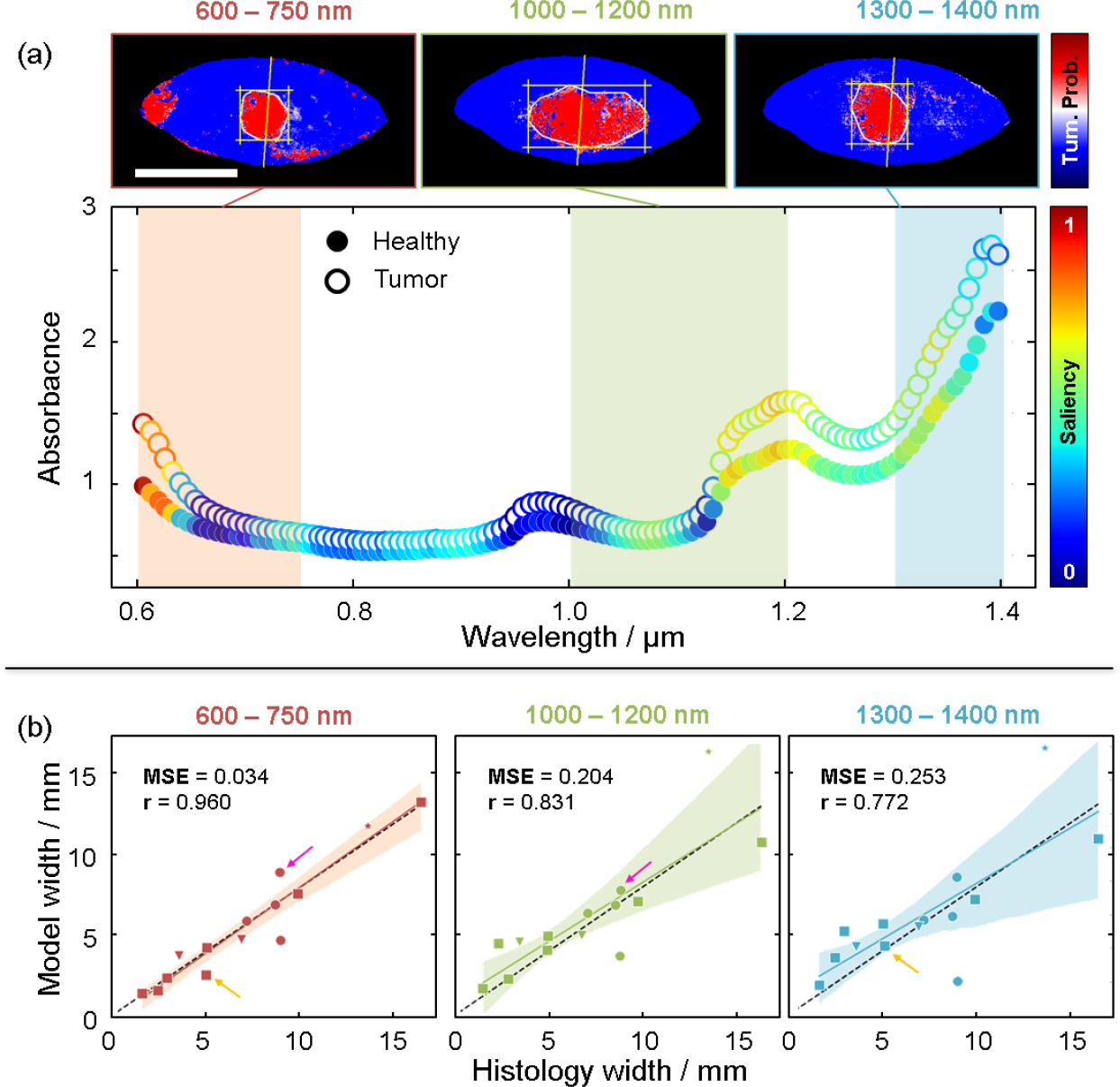
(a) Spectra taken from a BCC sample where the healthy tissue (solid markers) and tumor (hollow markers) are plotted with color-coding representing the saliency between 0 (blue) and 1 (red). Prediction maps are generated by reducing the spectral ranges used for the MCR-ALS generation of spectral components into three ranges: 600 nm – 750 nm (red shaded area), 1000 nm – 1200 nm (green shaded area), and 1300 nm – 1400 nm (blue shaded area). (b) Correlation plots for all tumors generated from prediction maps using the different spectral ranges where the spectral range between 600 nm – 750 nm yields an overall improved correlation. The colored arrows indicate samples that actually improve in the model prediction width in the other two spectral ranges, although the overall correlation is worse.

### Impact on model by using different spectral ranges

To assess the importance of different spectral ranges when classifying the pixels, we calculated the *saliency* (see Methods), which is a measure of how the output changes when the different input features are subjected to perturbations. The larger change in output for an input perturbation, the more importance, and higher saliency, the input has. Figure 6a shows a representative example of a BCC tumor sample where the spectra of healthy tissue and tumor are plotted with each data point color coded according to the saliency. The saliency is, naturally, the same for each spectral channel for the healthy tissue and tumor pixels. Since this is a binary classification task, the same features that are important to classify a pixel as healthy are the same with the ones being important for classifying a pixel as tumor.

The spectral ranges that differ the most between tumor and healthy pixels generally seem to have the largest saliency. The saliency peaks in different spectral ranges depending on the tumor (see SI Note VI for all tumors), which is consistent with the results obtained in Figure 2. Considering this, we trained the three models again, but this time only using selected spectral ranges: 600 nm – 750 nm, 1000 nm – 1200 nm and 1300 nm – 1400 nm. The three images in Figure 7a show how the resulting prediction maps obtaining the tumor borders for this BCC tumor change depending on the spectral range used by the MLP (see SI Figure S9-11 for all tumors). Figure 6b shows how the correlation between the histology tumor width and model tumor widths change for all tumors depending on the spectral range used. While the 600 nm – 750 nm spectral range yields an overall improved fit, demonstrated by a higher correlation coefficient and lower MSE, we note that this is again not the case for all tumors, which we show with the colored arrows that highlight some tumors that in fact improve when different spectral range are used. It is clear that the models pick up some different features depending on the spectral ranges and that the features are combined when using the full range. However, it is also apparent that each of the ranges contains enough information to naïvely find the tumors.

**Figure 7.**
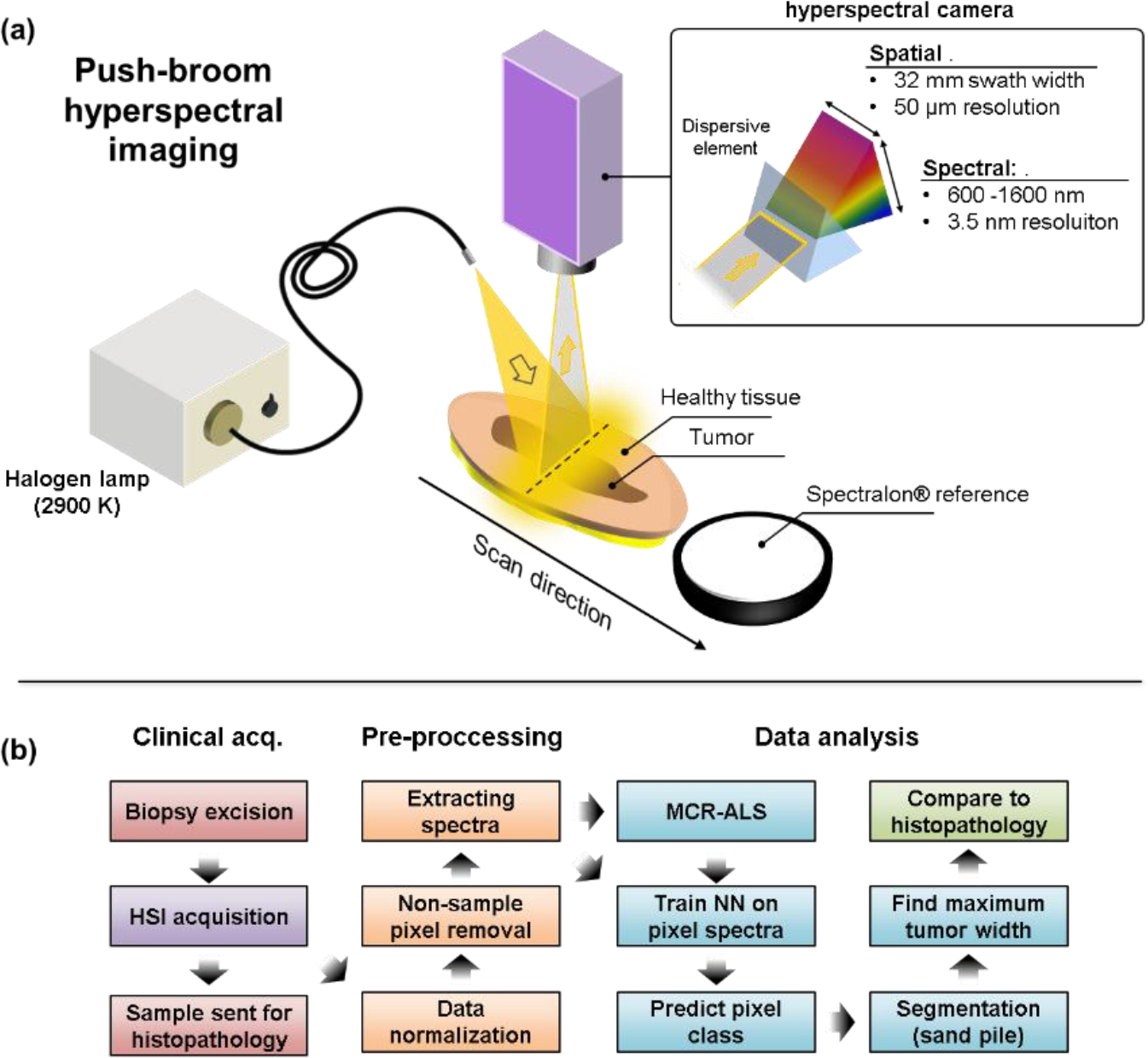
(a) Schematic demonstrating the spatial scanning hyperspectral measurement geometry in which a white incandescent lamp is used to illuminate the sample surface. A line is thereafter imaged, via a slit, onto an area detector. Prior to reaching the detector, the light is dispersed by a prism at each point along the line such that one dimension of the detector area captures the spectral information, while the other dimension captures the spatial information. By scanning the entire system across a surface, a hyperspectral image is obtained. A white reference placed at the same height as the sample is also scanned in every measurement. (b) Flow chart demonstrating the procedure going from acquisition of data from patient using HSI, through pre-processing of the data and finally to the data analysis.

Until now we have demonstrated that both the type of model used to generate a prediction map, and the spectral range used as a foundation for the pixel classifier, will yield different results depending on the tumor type. As such, there are several parameters that can be tweaked in order to optimize the algorithm for a particular tumor. While all possible parameters for each tumor type are far too many to be explored at this stage, we demonstrate that tweaking some parameters in regard to both model type and spectral range can significantly change the overall correlation and MSE of all tumors (see SI Figure S3), which points to a variation of spectral features for different tumors being captured across the broad spectral range that is used.

## Discussion

A tumor, by definition, constitutes a deviation in the molecular composition from healthy tissue. Our approach to skin tumor segmentation is therefore rooted in the fact that a change in the atomic or molecular composition translates into a change in the spectroscopic signature. ^36^ The limitation, however, is whether the spectroscopic change can be detected, which depends on both instrument capability and the molecular complexity of the examined tissue. While the molecular composition of healthy tissue is extremely complex and molecularly heterogeneous, ^37^ a tumor adds an unknown degree of molecular complexity. Additionally, since tissue properties are to an extent unique to each individual, it becomes challenging to develop one method accounting for all irregularities.^38^ This work presents a solution focused on maximizing the amount of spectral information extracted, which in turn drives the analysis to identify spectral disparities between healthy tissue and tumor within the same individual.

### Addressing a clinical need

A recent review addressing the challenges with employing machine learning for medical imaging highlights the lack of real clinical progress despite an increasing number of algorithms emerging aimed to facilitate more efficient diagnoses.^39^ The need for labelled ground truth images in CNN-based skin tumor delineation, for example, is in reality unrealistic for clinical implementation, which is not due to the need for histopathology, but also the potential systematic bias tied to the person doing the labeling.^40^ This work is derived from the need to improve current diagnostic practice, which is why we develop an approach not relying on labeled images.

### Circumventing dataset bias

A common hurdle for machine learning implementation is dataset bias, where the training data is not representative of the target population.^41^ The training data generated in our model certainly varies from patient to patient, however, this variation does not impede the performance. This is because the model does not identify spectral features that are similar between patients, but rather identifies how the spectrum representing tumor deviates from healthy tissue within the same patient. The training data is therefore only applicable to the patient in which it is acquired, which may seem limiting. However, our fundamentally different approach automatically accounts for inter-individual variations,^38^ which in fact increases its clinical applicability. Moreover, it may even find some utility for emerging precision diagnostics.^42^

### Spatial vs. spectral features for segmentation

In a recent review on different multispectral and hyperspectral spectroscopic approaches to various medical applications shows that most operate within the visible to near infra-red spectral range (400-1000 nm), with the number of spectral channels up to a 124.^43^ Present work, to the best of our knowledge, employs the broadest spectral range (600-1400 nm) with the most spectral channels (235). Although the novelty of this work hinges on the fact that the spectral features, that are more closely tied to the molecular profile of the sample, drive the machine learning and segmentation algorithms, we stress that an abundance of spectral information provides additional opportunity in the spectral feature extraction. A recent report compared using spatial and spectral features for determining melanoma skin tumor borders,^44^ although the spectral information was limited to only three channels in an RGB camera. Nonetheless, it was concluded that using the spectral information yielded the best results. With a broader and more resolved spectral range used in this work, more subtle spectral features can be identified, which should contribute to better differentiating between healthy tissue and tumor.

### Adaptability of the ML algorithm

Besides the abundance of spectral information available for the analysis, the ML algorithm has several parameters that can be tweaked to optimize the accuracy of the tumor border determination. The large number of parameter settings yielding satisfying performance (SI Figure S3) indicates robustness of the algorithm. The fact that tumor width correlation to histopathology changes for different tumors using the same set of parameters, and can be improved by changing the spectral range used, is actually encouraging. This is essentially a confirmation that we to some degree are not only able to probe the expected changes in molecular profile between different tumors and patients, but adapt the algorithm to account for these changes. Some degree of consistency is certainly expected between skin tumors of the same type, however, we can at this stage not draw any conclusions in regard to this due to the limited data set. We recognize that in order to draw such a conclusion requires a substantial amount of data, which may even require a tumor classification in relation to patient health data relating to tissue properties (i.e. Fitzpatrick skin type, age, smoking, etc.). A HSI system capable of generating an abundance of spectral information, coupled with a machine learning algorithm driven by spectral features, can better account for an increasing number of variables required for classification.

### Clinical impact

A careful and correct delineation of skin tumors prior to surgery would improve patient care by limiting the risk of non-radical excisions and subsequent re-operations, as well as sparing healthy tissue. In this study, hyperspectral imaging data from different skin tumor types was analyzed using artificial neural networks to visualize the extent of the tumors with a clear unbiased distinction from healthy skin without data input about the final diagnosis. Since the presented skin tumor delineation does not require any input of the actual border or any spatial features, it has a more realistic clinical implementation once some degree of reliability compared to histopathology can be determined through an extended clinical study. The only manual step of identifying the center of the tumor is a cheap and easy task so no clinical relevance is lost. Therefore, efforts to automate this step as well are postponed until more data is available. Within the framework of this study, a diagnosis of the tumor type based on spectral features is not possible due to the limited sample size, although previous studies suggests that classifying tumor type from regular dermoscopic images is possible. ^45^ The capability of the presented HSI method to easily generate an abundance of even more spectrally detailed data is therefore promising for current methods to be used for classification as well.

## Conclusion

This study demonstrates the utility of extracting detailed spectral information from suspected skin tumors and surrounding healthy tissue using HSI and ML for determining skin tumor borders. Based on the notion that there must be a difference in the molecular composition between healthy tissue and tumor, we demonstrate how only a few spectra from sample regions representing healthy and skin tumor can be used to determine the skin tumor border. By tuning the parameters of the machine learning algorithm, as well as the spectral content that is used, we highlight the adaptability of our approach to skin tumors in different individuals, as well as types. The determination of the skin tumor border is driven by a comparison to healthy tissue in the same individual, and the fact that the results change depending on the spectral range that is used demonstrates that we not only detect inter-individual variability in the spectral content, but can adapt the analysis to account for it. Current work not only provides insight into the potential use for HSI and spectroscopically-guided machine learning to be employed in emerging skin tumor precision diagnostic, but may also have an impact on current skin tumor diagnostic practice with increased likelihood of radical excision while minimizing removal of healthy tissue.

## Data Availability

The hyperspectral imaging data used in this work, as well as code used to generate the results, can be available upon request.

## Author contributions

E.A. developed the analysis method under the supervision of V.O., C.T. and P.E. J.H. conducted the measurements and oversaw patients together with M.S, B.S. and J.H.-P. under the supervision of B.P., M.M. and A.M. A.P.-L. conducted the histopathological analysis and consulted in interpretation of data. E.A., V.O., M.M. and A.M conceptualized and designed the study together, and wrote the manuscript. All authors revised the final version of the manuscript.

## Competing interests

The authors declare no competing interests.

## Funding

This study was supported by the Swedish Government Grant for Clinical Research (ALF), Skåne University Hospital (SUS) Research Grants, Lund University grant for Research Infrastructure. Skåne County Council Research Grants, Sjögrens Stiftelse. AM acknowledges Lund University Medical Faculty (F2022/1896) and Carmen and Bertil Regnér Foundation (2022-00083). VO gratefully acknowledges the support of the US National Institutes of Health (USPHS grant R01HL119102) and Crafoordska Stiftelsen grant 20200859. This work was supported by a grant from the Knut and Alice Wallenberg Foundation to SciLifeLab for research in Data-driven Life Science, DDLS (KAW 2020.0239). This work was partially supported by the Wallenberg AI, Autonomous Systems and Software Program (WASP) funded by the Knut and Alice Wallenberg Foundation.

## Methods

### Ethics

The study was approved by the Ethics Committee at Lund University, Sweden (2022-04900-02). The research adhered to the tenets of the Declaration of Helsinki as amended in 2013. Prior to surgery, all the patients participating in the study were given verbal and written information about the study and the voluntary nature of participation. All patients gave their informed written consent.

### Subjects

21 patients were included in the study with a total of 22 skin tumors suspected of melanoma, basal cell carcinoma (BCC), or squamous cell carcinoma (SCC). One of the patients had 2 skin tumors. After histopathological diagnosis, three samples were excluded from analysis since diagnosis revealed either sun damaged skin or bluish colored skin with no measurable lesion sizes. On the Fitzpatrick scale, all samples ranged from skin type I (pale complexion) to type VI (dark complexion), with a vast majority being of skin type II. See Table 1 for details on the samples included for analysis in the study. Ages are reported as a range spanning 10 years (i.e. 60’s includes all ages between 60-69 years).

**Table 1:** Patient data.

| Lesion (patient) no. | Sex | Age | Fitzpatrick skin type (I-VI) | Diagnosis |
| --- | --- | --- | --- | --- |
| 1 (1) | Male | 60's | I | Malignant melanoma, in situ |
| 2 (2) | Male | 50's | II | Dysplastic nevus |
| 3 (3) | Male | 60's | II | Fibrohistiocytoma |
| 4 (4) | Male | 60's | II | Basal cell carcinoma |
| 5 (5) | Male | 50's | N/I | Papilloma |
| 6 (6) | Female | 40's | II | Dysplastic nevus |
| 7 (7) | Male | 80's | II | Squamous cell carcinoma |
| 8 (8) | Male | 2's | IV | Blue nevus |
| 9 (9) | Female | 40's | II | Nevus |
| 10 (10) | Female | 70's | II | Basal cell carcinoma |
| 11 (11) | Female | 60's | II | Malignant melanoma, in situ |
| 12 (12) | Female | 70's | II | Basal cell carcinoma |
| 13 (13) | Female | 60's | II | Squamous cell carcinoma |
| 14 (14) | Male | 60's | II | Nevus |
| 15 (15) | Male | 60's | II | Nevus and blue nevus |
| 16 (16) | Female | 60's | II | Basal cell carcinoma |
| 17 (17) | Male | 40's | II | Malignant melanoma, invasive |
| 18 (17) | Male | 40's | II | Nevus |
| 19 (18) | Male | 80's | II | Malignant melanoma, invasive |

### Examination and measurement procedure

All suspected tumors were excised under local anesthesia by experienced dermatologists, using standard local excision recommendations. They were then placed in isotonic saline for a short period of time before being imaged with HSI. After imaging, the tumors were placed in formaldehyde and sent for histopathological examination.

### Hyperspectral imaging

A hyperspectral image can be generated in a few different ways. For a detailed description of the different HSI techniques see.^26^ However, we describe here the essence of what separates the various techniques. HSI aims to acquire information in both the spectral and spatial domains, which is achieved over a period of time (temporal domain). This establishes a three-way trade-off in instrument design, where different methods maximize the information gained in one domain over another. Scanning the spectral domain in time ensures integrity in the spatial domain at the expense of integrity in the spectral domain. Scanning in the spatial domain, ensures spectral over spatial integrity. Snapshot HSI methods are emerging where a hyperspectral image is generated instantaneously, although this comes at the expense of sacrificing information in the both the spectral and spatial domains. One technique is not better than another, rather, the HSI approach should be suited for the type of sample that is imaged, and the measurement domain where data integrity is most critical.

As will be emphasized throughout this work, spectral information will be of primary importance, which is why we opt for a so-called =spatial scanning’ HSI technique. These techniques operate by imaging a single column (detection line) of the object onto a sensor array after having passed through some spectrally dispersive medium. At a cost of reducing the image to only one spatial dimension, complete spectral information is acquired for all points along the measured line. Thereafter, either the sample or the camera slightly moves to capture information from an adjacent line of the object (Figure 7a). The hyperspectral camera was custom-built in the HySpex model series (Norsk Elektro Optikk, Oslo, Norway). A halogen lamp with a peak temperature of 2900 K was used for excitation, which yielded a broad emission profile with a peak wavelength at 1000 nm. The camera uses a combination of a Si and InGaAs detectors, which together with the excitation effectively enables VIS-NIR-SWIR spectral sensitivity in the range between 600 nm – 1700 nm. The spectral bandwidth is divided into 322 spectral bands, yielding a spectral resolution of approximately 3.5 nm. An objective with an angular field-of-view of 16° and working distance of 75 mm was used resulting in an effective swath width of 32 mm. In combination with the sensor having 640 pixels, a spatial resolution of 50 μm is obtained. The exposure time of the camera ranges between 10-90 ms, which means that a sample 5 cm in length takes up to 90 seconds to measure.

### Reference measurements, normalization and absorbance calculations

Prior to imaging, an internal shutter is inserted in front of the camera and a signal is recorded at the specified exposure time which is thereafter subtracted from all recorded images. This accounts primarily for the read-out noise generated by camera. To account for the ambient background light, a measurement is made without the excitation light source on (I_Bkg_(λ)). A white reference signal is obtained by scanning across a Spectralon® white reference (WS1, Ocean Optics) that is placed in the scanning path after the sample as demonstrated in Figure 7a. Thus, the white reference and the sample measurement are obtained in the same measurement. A white reference spectrum (I_Ref_(λ)) is obtained in every pixel along the detection line to account for potential intensity variation perpendicular to the scan path. Thereafter, the spectrum (I(λ)) in every pixel in the image is normalized against the corresponding white reference spectrum obtained in the same horizontal line to ensure that the correct intensity from the light source is used. Thereafter, absorbance as a function of wavelength (A(λ)) can be calculated according to,

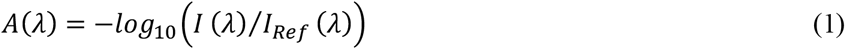

### Pre-processing: Non-sample pixel removal & spectra extraction

Once the data has been normalized according to Eq. (1) above, pixels not representing the sample needed to be removed. We based this analysis step on the notion that the most significant spectral variation in the entire image is between the sample and non-sample (background). We performed a principal component analysis (PCA) on all the pixel spectra. Using only the first two principal components, we clustered the pixels using HDBscan^46,47^ with *min_cluster_size* = 500 and otherwise the standard parameters. Smaller clusters were merged until two clusters remained. Since the hyperspectral images were produced with the sample centered, the background cluster could automatically be identified as the one containing the corner pixels, and the remaining central cluster as the sample. Thereafter, the absorbance spectrum for all pixels represented by sample were extracted with their image coordinates tagged.

### Multivariate Curve Resolution – Alternating Least Squares

To aid in visualization of the high-dimensional spectral data, MCR-ALS (multivariate curve resolution – alternating least squares) analysis^30^ was applied to reduce dimensionality in an intuitive manner. All tumor hyperspectral images were merged into one composite hyperspectral image, then decomposed into a sum of eight spectra-like components, with their contributions to each pixel in the image being all non-negative. This spatial pattern was then defined by just eight numbers that may be visualized directly but also present an alternative to using the full spectra in subsequent analysis steps. In essence, this step generates a set of spectra that together capture the most common spectral features of all tumors combined. With this information, one can assess the extent to which each pixel spectrum for a particular tumor can be represented by each of these spectra, which assists in the visualization of potentially deviating tissue properties (i.e., tumor).

### Computational framework

Machine learning is a type of artificial intelligence that uses training data to automatically improve computer algorithms, in order to make predictions. A model, for example an artificial neural network (ANN) or a support-vector machine (SVM), is used to train the system on a set of limited data, allowing the system to generalize to, and predict outcome of, unseen data.

We have devised a computational framework which predicts superficial shape and size of a skin tumor from a hyperspectral image. The structure of the data analysis framework, together with the data acquisition and pre-processing, is shown as a flowchart in Figure 7b. An ANN is trained on the spectra of individual pixels from a selection of pixels with known tumor/non-tumor state. We emphasize that no spatial features are needed or used to train the ML algorithm, rather, it is the spectral features that are used. The trained ANN is then used to predict whether the rest of the pixels in the image belong to either tumor or non-tumor (presumed healthy tissue) regions. This generates a prediction map for the tumor which serves as input for a two-step segmentation algorithm (described below).

The total framework, consisting of a combination of an ANN and a segmentation algorithm, predicts the size and delineation of the tumor and does not rely on any prior knowledge of tumor borders. It only uses the information contained in one single HS image. Thus, patient variability, e.g., skin complexion, does not provide bias to the predictions. Neither is the framework restricted to certain types of tumors since only the spectral variation between the same patient’s healthy tissue and tumor is utilized. All details are found in the supporting information (SI), while we provide below only the necessary aspects needed for understanding the presented results.

### Machine learning

#### Artificial neural networks

We developed three different machine learning models to classify individual pixels as either “healthy tissue” or “tumor” solely based on spectral information. As ANNs, we used a multilayer perceptron (MLP) trained on all spectral components between 600 nm and 1400 nm (235 channels), an MLP trained on the MCR-ALS decomposition (eight components; see method) of the spectra, and a one-dimensional convolutional neural network (CNN) trained on all the spectral components between 600 nm and 1400 nm (see SI Note I and Figure S4 for model details). All models were developed using Python 3.7 and Tensorflow 1.14.

A separate model instance was trained for each tumor sample. We selected pixels from the center of the tumor and from regions of the sample likely representing healthy tissue and then used their spectra as training data for supervised training. To mitigate any artifacts resulting from the scanning direction of the camera, we chose two areas of healthy pixels, one from each side of the tumor along the scanning axis. The sizes of the two healthy areas were chosen so that they in total contain approximately as many pixels as the tumor area to ensure balanced training datasets (see SI Figure S1). An ensemble of classifiers was trained for each model and sample using *K*-fold cross splitting with *K=*5. The trained ensembles predict the probability of each pixel representing healthy tissue or tumor, where the average of the predicted value for each ensemble member is used as the ensemble prediction. A threshold of 0.5 was used to differentiate between pixels classified as healthy tissue and tumor (see SI Note I for further information).

#### Saliency

The saliency was used as a feature importance measure for each feature *k*. The saliency for a feature is defined as the change of the output value as response to a change in the input feature, averaged over all patterns,

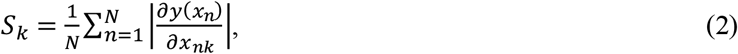

where *y* is the output value given input pattern ***x***_*n*_ and *N* is the number of patterns.

#### Segmentation

An active contour segmentation algorithm^35^ was used to determine the tumor delineation. In the algorithm, an active contour, here referred to as a *rope*, tracks the outline of an object in an image by minimizing an energy function. The position of the rope can parametrically be described as *r*(*p*) = (*x*(*p*), *y*(*p*)) with *r*(0) = *r*(1)meaning that the rope is a closed loop. The general energy function of the rope consists of internal and external energy contributions as well as a constraint contribution.

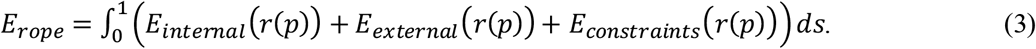

Here, we use

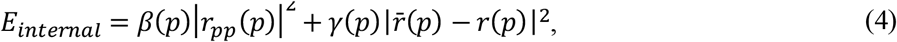

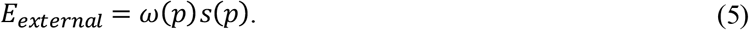

and

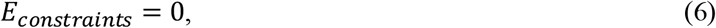

where *α*(*p*), *β*(*p*) and *ω*(*p*) are hyper-parameters and ***s*** is the sand-pile function described below (see SI Note II for computational details).

The sand-pile function *s*(*p*) replaces the pixel intensity of standard active contour segmentation in the external energy. A sand-pile is constructed on the prediction maps to give each pixel classified as tumor a spatial context, i.e., if a pixel is located in the center or edge of the tumor. Conceptually, sand-piles are constructed by pouring sand on the pixels classified as tumorous. As the pixels classified as healthy tissue act as sinks, a sand-pile will emerge on the tumor. The height of the resulting sand-pile is greater in the center of the tumor. The rope in the active contour algorithm is then tightened around the sand-pile. We refer the reader to SI Note II for supplementary mathematical and computational details.

#### Evaluation (statistical) methods

To evaluate the models, the histopathological measurements of the tumor widths were compared to the model predicted widths. The mean subtracted Pearson correlation was calculated, as well as the relative mean squared error (MSE), defined by

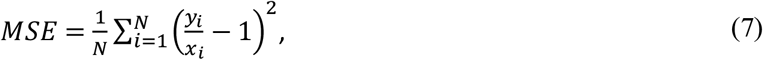

where *x*_*i*_ is the histopathological width and *y*_*i*_ is the model predicted width for sample *i* and *N* is the number of samples. We consider the model predictions with higher Pearson correlation coefficient and lower MSE to be the better ones.

## Notes

### Competing Interest Statement

The authors have declared no competing interest.

### Funding Statement

This study was supported by the Swedish Government Grant for Clinical Research (ALF), Skane University Hospital (SUS) Research Grants, Lund University grant for Research Infrastructure. Skane County Council Research Grants, Sjogrens Stiftelse. AM acknowledges Lund University Medical Faculty (F2022/1896) and Carmen and Bertil Regner Foundation (2022-00083). VO gratefully acknowledges the support of the US National Institutes of Health (USPHS grant R01HL119102) and Crafoordska Stiftelsen grant 20200859. This work was supported by a grant from the Knut and Alice Wallenberg Foundation to SciLifeLab for research in Data-driven Life Science, DDLS (KAW 2020.0239). This work was partially supported by the Wallenberg AI, Autonomous Systems and Software Program (WASP) funded by the Knut and Alice Wallenberg Foundation.

### Author Declarations

The study was approved by the Ethics Committee at Lund University, Sweden (2022-04900-02).

## References

1. Sung, H. et al. Global Cancer Statistics 2020: GLOBOCAN Estimates of Incidence and Mortality Worldwide for 36 Cancers in 185 Countries. CA. Cancer J. Clin. 71, 209–249 (2021).

2. Neville, J. A., Welch, E. & Leffell, D. J. Management of nonmelanoma skin cancer in 2007. Nat. Clin. Pract. Oncol. 4, 462–469 (2007).

3. Ellison, P. M., Zitelli, J. A. & Brodland, D. G. Mohs micrographic surgery for melanoma: A prospective multicenter study. J. Am. Acad. Dermatol. 81, 767–774 (2019).

4. Siegel, R. L., Miller, K. D. & Jemal, A. Cancer statistics, 2019. CA. Cancer J. Clin. 69, 7–34 (2019).

5. Liopyris, K., Gregoriou, S., Dias, J. & Stratigos, A. J. Artificial Intelligence in Dermatology: Challenges and Perspectives. Dermatol. Ther. (Heidelb). 12, 2637–2651 (2022).

6. Beltrami, E. J. et al. Artificial intelligence in the detection of skin cancer. J. Am. Acad. Dermatol. 87, 1336–1342 (2022).

7. Smak Gregoor, A. M. et al. An artificial intelligence based app for skin cancer detection evaluated in a population based setting. npj Digit. Med. 6, 1–8 (2023).

8. Soenksen, L. R. et al. Using deep learning for dermatologist-level detection of suspiUsing deep learning for dermatologist-level detection of suspicious pigmented skin lesions from wide-field imagescious pigmented skin lesions from wide-field images. Sci. Transl. Med. 13, 1–13 (2021).

9. Cancer Council Australia Keratinocyte Cancers Guideline Working Party. Clinical practice guidelines for keratinocyte cancer. Sydney: Cancer Council Australia. [Version URL: https://wiki.cancer.org.au/australiawiki/index.php?oldid=213931, xcited 2023 Jun 9].

10. Cancer Council Australia Melanoma Guidelines Working Party. Clinical practice guidelines for the diagnosis and management of melanoma. Sydney: Melanoma Institute Australia. [Version URL: https://wiki.cancer.org.au/australiawiki/index.php?oldid=215123, cit.

11. Lisa, A. V. E. et al. Outpatient Nonmelanoma Skin Cancer Excision and Reconstruction: A Clinical, Economical, and Patient Perception Analysis. Plast. Reconstr. Surg. - Glob. Open 10, E3925 (2022).

12. Greiff, L., Skogvall-Svensson, I., Carneiro, A. & Hafström, A. Non-radical primary diagnostic biopsies affect survival in cutaneous head and neck melanoma. Acta Otolaryngol. 141, 309–319 (2021).

13. Mirikharaji, Z. et al. A survey on deep learning for skin lesion segmentation. Med. Image Anal. 88, 102863 (2023).

14. Navarrete-Dechent, C., Liopyris, K. & Marchetti, M. A. Multiclass Artificial Intelligence in Dermatology: Progress but Still Room for Improvement. J. Invest. Dermatol. 141, 1325–1328 (2021).

15. Young, A. T. et al. Stress testing reveals gaps in clinic readiness of image-based diagnostic artificial intelligence models. npj Digit. Med. 4, (2021).

16. Goyal, M., Knackstedt, T., Yan, S. & Hassanpour, S. Artificial intelligence-based image classification methods for diagnosis of skin cancer: Challenges and opportunities. Comput. Biol. Med. 127, 104065 (2020).

17. Cullell-Dalmau, M., Noé, S., Otero-Viñas, M., Meić, I. & Manzo, C. Convolutional Neural Network for Skin Lesion Classification: Understanding the Fundamentals Through Hands-On Learning. Front. Med. 8, 1–8 (2021).

18. Papageorgiou, V. et al. The limitations of dermoscopy: false-positive and false-negative tumours. J. Eur. Acad. Dermatology Venereol. 32, 879–888 (2018).

19. Dinnes, J. et al. Dermoscopy, with and without visual inspection, for diagnosing melanoma in adults (Review). Cochrane Database Syst. Rev. CD011902 (2018) doi:10.1002/14651858.CD011902.pub2. http://www.cochranelibrary.com.

20. Ali, A. R., Li, J., Yang, G. & O’Shea, S. J. A machine learning approach to automatic detection of irregularity in skin lesion border using dermoscopic images. PeerJ Comput. Sci. 6, 1–35 (2020).

21. Hwang, Y. N., Seo, M. J. & Kim, S. M. A Segmentation of Melanocytic Skin Lesions in Dermoscopic and Standard Images Using a Hybrid Two-Stage Approach. Biomed Res. Int. 2021, 5562801 (2021).

22. Van Molle, P. et al. Dermatologist versus artificial intelligence confidence in dermoscopy diagnosis: Complementary information that may affect decision-making. Exp. Dermatol. 1–8 (2023) doi:10.1111/exd.14892.

23. Vestergaard, M. E., Macaskill, P., Holt, P. E. & Menzies, S. W. Dermoscopy compared with naked eye examination for the diagnosis of primary melanoma: A meta-analysis of studies performed in a clinical setting. Br. J. Dermatol. 159, 669–676 (2008).

24. Haggenmüller, S. et al. Skin cancer classification via convolutional neural networks: systematic review of studies involving human experts. Eur. J. Cancer 156, 202–216 (2021).

25. Pertzborn, D. et al. Intraoperative Assessment of Tumor Margins in Tissue Sections with Hyperspectral Imaging and Machine Learning. Cancers (Basel). 15, (2023).

26. Lu, G. & Fei, B. Medical hyperspectral imaging: a review. J. Biomed. Opt. 19, 010901 (2014).

27. Lindholm, V. et al. Differentiating Malignant from Benign Pigmented or Non-Pigmented Skin Tumours—A Pilot Study on 3D Hyperspectral Imaging of Complex Skin Surfaces and Convolutional Neural Networks. J. Clin. Med. 11, (2022).

28. Johansen, T. H. et al. Recent advances in hyperspectral imaging for melanoma detection. Wiley Interdisciplinary Reviews: Computational Statistics vol. 12 1–17 at 10.1002/wics.1465 (2020).

29. Hu, L., Luo, X. & Wei, Y. Hyperspectral Image Classification of Convolutional Neural Network Combined with Valuable Samples. J. Phys. Conf. Ser. 1549, (2020).

30. Jaumot, J., de Juan, A. & Tauler, R. MCR-ALS GUI 2.0: New features and applications. Chemom. Intell. Lab. Syst. 140, 1–12 (2015).

31. Ronnberger, O., Fischer, P. & Brox, T. U-Net: Convolutional Networks for Biomedical Image Segmentation. arXiv:1505.04597 (2015) doi:10.1109/ACCESS.2021.3053408.

32. Leon, R. et al. Non-invasive skin cancer diagnosis using hyperspectral imaging for in-situ clinical support. J. Clin. Med. 9, (2020).

33. Li, W., Joseph Raj, A. N., Tjahjadi, T. & Zhuang, Z. Digital hair removal by deep learning for skin lesion segmentation. Pattern Recognit. 117, 107994 (2021).

34. Kim, B., Kim, Hy., Kim, K., Kim, S. & Kim, J. Learning Not to Learn: Training Deep Neural Networks With Biased Data. in IEEE/CVF Conference on Computer Vision and Pattern Recognition (CVPR) 9004–9012 (2019).

35. Kass, M., Witkin, A. & Terzopoulos, D. Snakes: Active Contour Models. Int. J. Comput. Vis. 321–331 (1988) doi:10.1016/j.procs.2018.10.231.

36. Svanberg, S. Atomic and Molecular Spectroscopy Sune Svanberg Basic Aspects and Practical Applications. (2022).

37. Lister, T., Wright, P. A. & Chappell, P. H. Optical properties of human skin. J. Biomed. Opt. 17, 0909011 (2012).

38. E., J., J., C., J., P. & P., S. Intra- and inter-individual variability in the mechanical properties of the human skin from in vivo measurements on 20 volunteers. Ski. Res. Technol. 23, 491–499 (2017).

39. Varoquaux, G. & Cheplygina, V. Machine learning for medical imaging: methodological failures and recommendations for the future. npj Digit. Med. 5, (2022).

40. Joskowicz, L., Cohen, D., Caplan, N. & Sosna, J. Inter-observer variability of manual contour delineation of structures in CT. Eur. Radiol. 29, 1391–1399 (2019).

41. Dockès, J., Varoquaux, G. & Poline, J.-B. Preventing dataset shift from breaking machine-learning biomarkers J er Introduction : Dataset Shift Breaks Learned A Primer on Machine Learning for Biomarkers. Gigascience 10, 1–11 (2021).

42. Bhattacharya, A. et al. Precision Diagnosis Of Melanoma And Other Skin Lesions From Digital Images. AMIA Jt. Summits Transl. Sci. proceedings. AMIA Jt. Summits Transl. Sci. 2017, 220–226 (2017).

43. Aloupogianni, E., Ishikawa, M., Kobayashi, N. & Obi, T. Hyperspectral and multispectral image processing for gross-level tumor detection in skin lesions: a systematic review. J. Biomed. Opt. 27, 1–28 (2022).

44. Annaby, M. H., Elwer, A. M., Rushdi, M. A. & Rasmy, M. E. M. Melanoma Detection Using Spatial and Spectral Analysis on Superpixel Graphs. J. Digit. Imaging 34, 162–181 (2021).

45. Esteva, A. et al. Dermatologist-level classification of skin cancer with deep neural networks. Nature 542, 115–118 (2017).

46. Campello, R. J. G. B., Moulavi, D. & Sander, J. Density-based clustering based on hierarchical density estimates. in Lecture Notes in Computer Science (including subseries Lecture Notes in Artificial Intelligence and Lecture Notes in Bioinformatics) vol. 7819 LNAI 160–172 (2013).

47. McInnes, L., Healy, J. & Astels, S. hdbscan: Hierarchical density based clustering. J. Open Source Softw. 2, 205 (2017).

