## supplemental information for "Facilitating clinically relevant skin tumor diagnostics with spectroscopy-driven machine learning"

### Supplementary Information for
Facilitating clinically relevant skin tumor diagnostics with spectroscopy-driven machine learning

Emil Andersson, Jenny Hult, Carl Troein,
Magne Stridh, Benjamin Sjögren, Agnes Pekar-Lukacs,
Julio Hernandez-Palacios, Patrik Edén, Bertil Persson,
Victor Olariu, Malin Malmsjö, Aboma Merdasa

### SI Note I: Machine Learning model details

#### Data description

The dataset consists of 18 samples. Table S1 lists the number of sample pixels (i.e. those pixels that were separated from the background pixels as described in Methods), and the number of pixels in the chosen training dots for tumor and healthy tissues respectively. Also the tumor widths measured by the pathologists are listed. The defined training dots are also displayed for all samples in Figure S1

**Table S1: Data properties.** The number of pixels each sample consists of, the number of pixels in the selected tumor and healthy tissue areas, and the samples' histopathological widths.

| Lesion no. | # sample pixels | # tumorous pixels | # healthy pixels | Histopathological width (mm) |
|---|---|---|---|---|
| 1 | 93363 | 709 | 754 | 6.89 |
| 2 | 210700 | 1961 | 2514 | 15.7 |
| 3 | 152591 | 1009 | 754 | 6.78 |
| 4 | 30534 | 197 | 226 | 7.83 |
| 6 | 27404 | 197 | 226 | 2.39 |
| 7 | 136424 | 1257 | 1226 | 13 |
| 8 | 174442 | 1009 | 1226 | 9.45 |
| 9 | 49739 | 1257 | 298 | 4.89 |
| 10 | 49468 | 1961 | 2018 | 3.45 |
| 11 | 149952 | 1257 | 1594 | 8.58 |
| 12 | 143629 | 1653 | 1594 | 6.76 |
| 13 | 46080 | 529 | 394 | 4.97 |
| 14 | 62715 | 709 | 506 | 2.86 |
| 15 | 18392 | 317 | 298 | 1.6 |
| 16 | 55359 | 709 | 754 | 6.61 |
| 17 | 112117 | 1653 | 1594 | 8.31 |
| 18 | 52377 | 613 | 754 | 4.84 |
| 19 | 145197 | 1961 | 2018 | 8.54 |

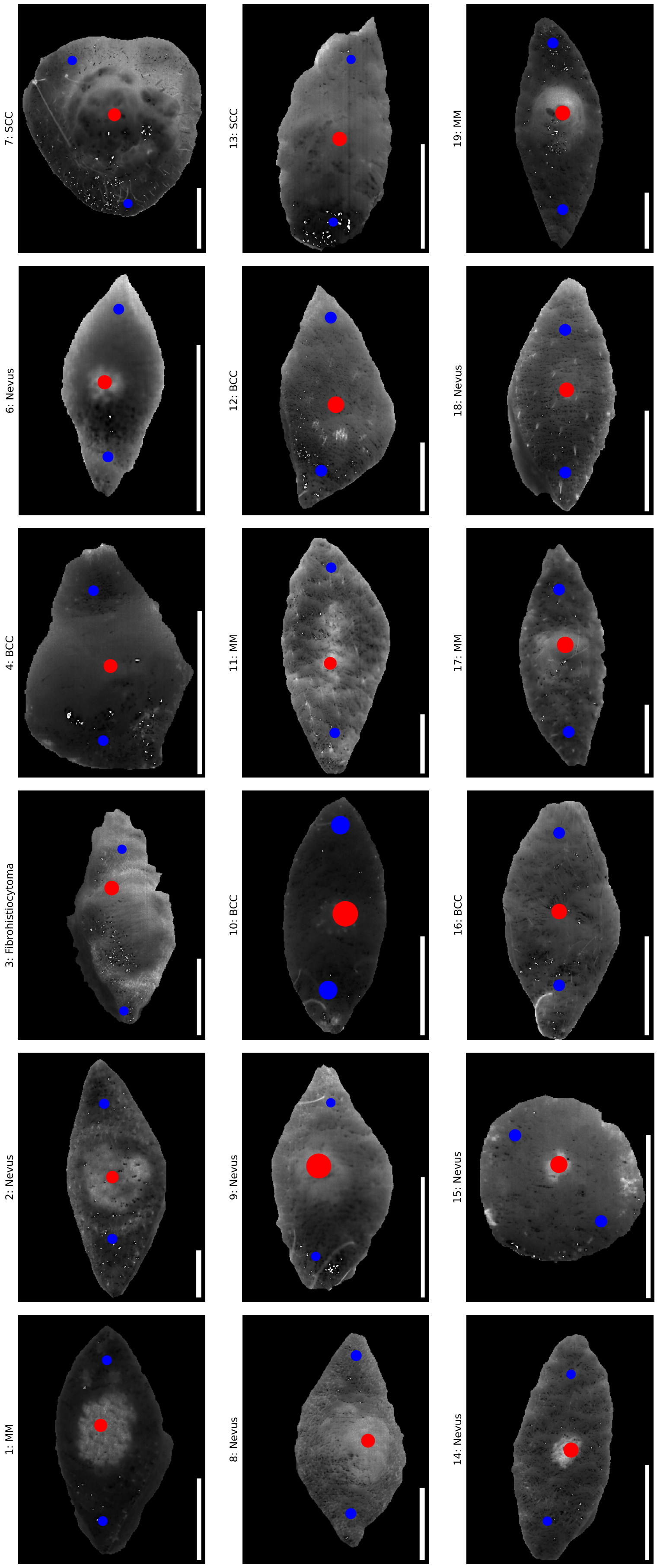

**Figure S1: Training regions**. The training regions used for each sample with blue representing healthy tissue and red tumor. The tumor samples are showned at 640 nm.

#### Training procedure

The hyperparameter search was conducted with a grid search where network architecture, learning rate and batch-size were changed, see the used values in Table S2. For the MLPs the hyperbolic tangent (tanh) was used as activation function, while rectified linear unit (ReLU) was used as activation function for the CNNs. All the models had one output node with the sigmoid function as output activation function. The binary cross-entropy was used as cost function and the models were optimized using Adam.[1] The pixels in the training dots (Table S1 and Figure 4a) were divided into 5 equally sized parts. One part each part was used for validation for the different members of the ensemble.

**Table S2: Model hyperparametes**

**(a) Grid search parameters.**

| Hyperparameter | Search values |
|---|---|
| Learning rate | $[0.025, 0.07, 0.01, 0.15]$ |
| Batch size | $[0.2, 0.5, 1.0]$ |

**(b) Network architectures.** The square brackets denotes an architecture where each item is a hidden layer. All convolutional filters and maxpoolings are 1-dimensional. $X$C denotes number of channels and $Y$S denotes stride $Y$.

| Model | Architecture |
|---|---|
| MLP,<br>MLP: MCR-ALS | [2], [8], [20], [100], [2,2], [8,8], [100,100], [2,2,2], [8, 8, 8] |
| CNN | [Conv(10, 10C, 10S), MaxPool(2, 2S), Conv(5, 20C, 1S), Dense(10)],<br>[Conv(10, 10C, 10S), MaxPool(2, 2S), Conv(5, 20C, 1S),<br>MaxPool(2, 2S), Conv(5, 20C, 1S), Dense(10)],<br>[Conv(10, 10C, 5S), MaxPool(2, 2S), Conv(5, 20C, 5S),<br>MaxPool(2, 2S), Conv(3, 20C, 2S), Dense(10)],<br>[Conv(20, 10C, 10S), MaxPool(2, 2S),<br>Conv(10, 20C, 5S), Dense(10)],<br>[Conv(5, 10C, 1S), Conv(10, 10C, 1S), MaxPool(2, 2S),<br>Conv(10, 20C, 1S), Dense(10)],<br>[Conv(20, 10C, 5S) MaxPool(2, 2S), Conv(10, 10C, 2S),<br>MaxPool(2, 2S), Conv(5, 10C, 1S), Dense(10)] |

#### Validation procedure

Model selection could not be performed the conventional way by picking the model with the best performance on the validation performance since most models achieved perfect accuracy (1.0) and loss (0) on both training and validation data (Figure S2). This is due to the simplicity of the classification of the blue and red regions in Figure 2a and b, which corresponds to the training areas (blue and red dots) in Figure 4a and Figure S1. However, the challenge of classifying tumor border pixels, which are outside these regions, remains.

Model validation at this stage is not feasible since we do not have labels for the border pixels. The only measured data we can validate the models against is the histopathological tumor widths. Hence, we first need to predict the tumor widths with all the potential models before model selection can be performed.

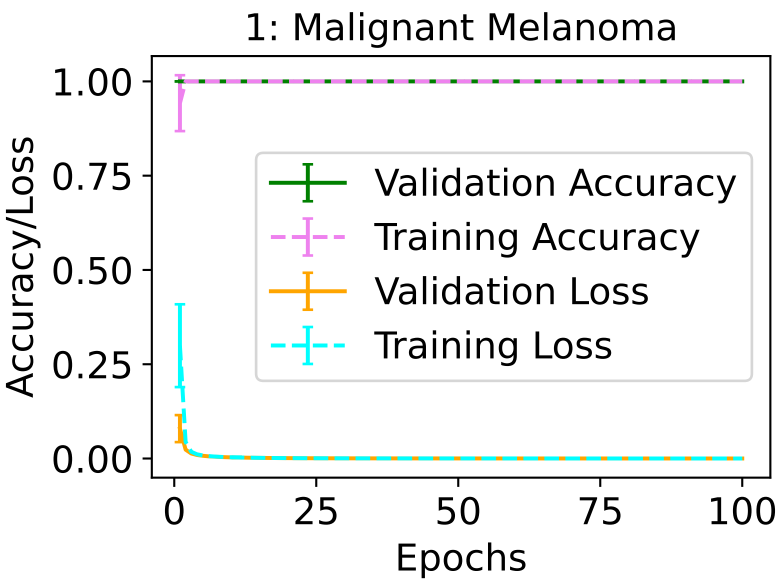

**Figure S2: Training and validation performance history.** Training and validation accuracy and loss history for a representative sample during training. The errorbars indicate the standard deviation.

The Pearson correlation coefficient and relative MSE (Metods) were calculated for each model and were plotted against each other (Figure S3). Each dot represents a model in the scatter plot. A model is considered to perform better the higher the correlation and the lower the MSE are, corresponding to the top-left corner of the plot. The models showed results for are the top performing model of each model type: MLP, CNN and MLP: MCR-ALS. The details of the chosen models are shown in Figure S4.

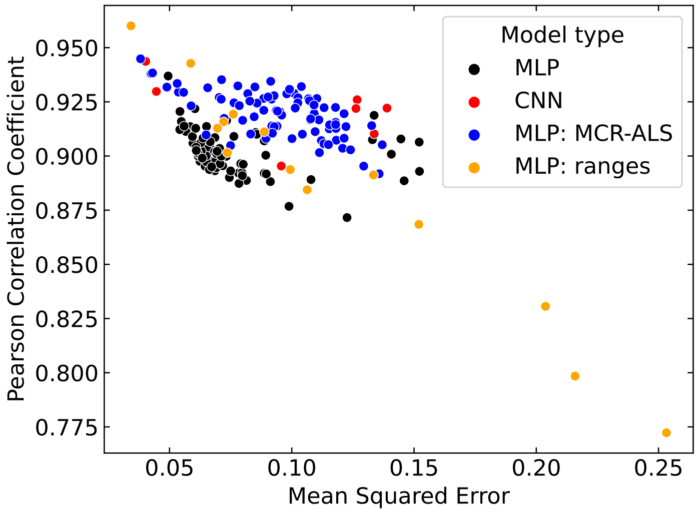

**Figure S3: Model validation.** Each dot represent a model from the grid search (Table S2) in the realative MSE - Pearsen Correlation Coefficient space. The color of the dots represent the different kinds of models. The orange orange "MLP: ranges" refer to the MLP trained of various sets of ranges. The top-left corner represents the lowest error and highest correlation.

### SI Note II: Computational details

#### Active Contour Segmentation

Computationally, the rope consists of a set of $n(t)$ points which are initially located along the sample border with one point per pixel. The position of the rope is iteratively updated where, at each time-step, each point is moved to the neighboring pixel which minimizes the energy function locally. The resolution of the rope is adaptive; the number of points comprising the rope changes. If the distance between two neighboring points of the rope gets more than 10 pixels apart, a new point is added between them. If two neighboring points get closer than 2 pixels apart, they are merged. During the first 30 steps, only the gravitational term contributes to the energy function to make sure that the rope does not get stuck on the sample border. The algorithm is terminated if the rope is identical after an update, or if a maximum number of updates has been made. The final position of the rope is considered as the tumor delineation. See Table S3 for the parameters used.

**Table S3: Parameters for the active contour algorithm.**

| Time step | $\beta$ | $\gamma$ | $\omega$ |
|---|---|---|---|
| $0 < t < 30$ | 0 | 10 | 0 |
| $30 \leq t < 500$ | 1 | 1 | 0.5 |

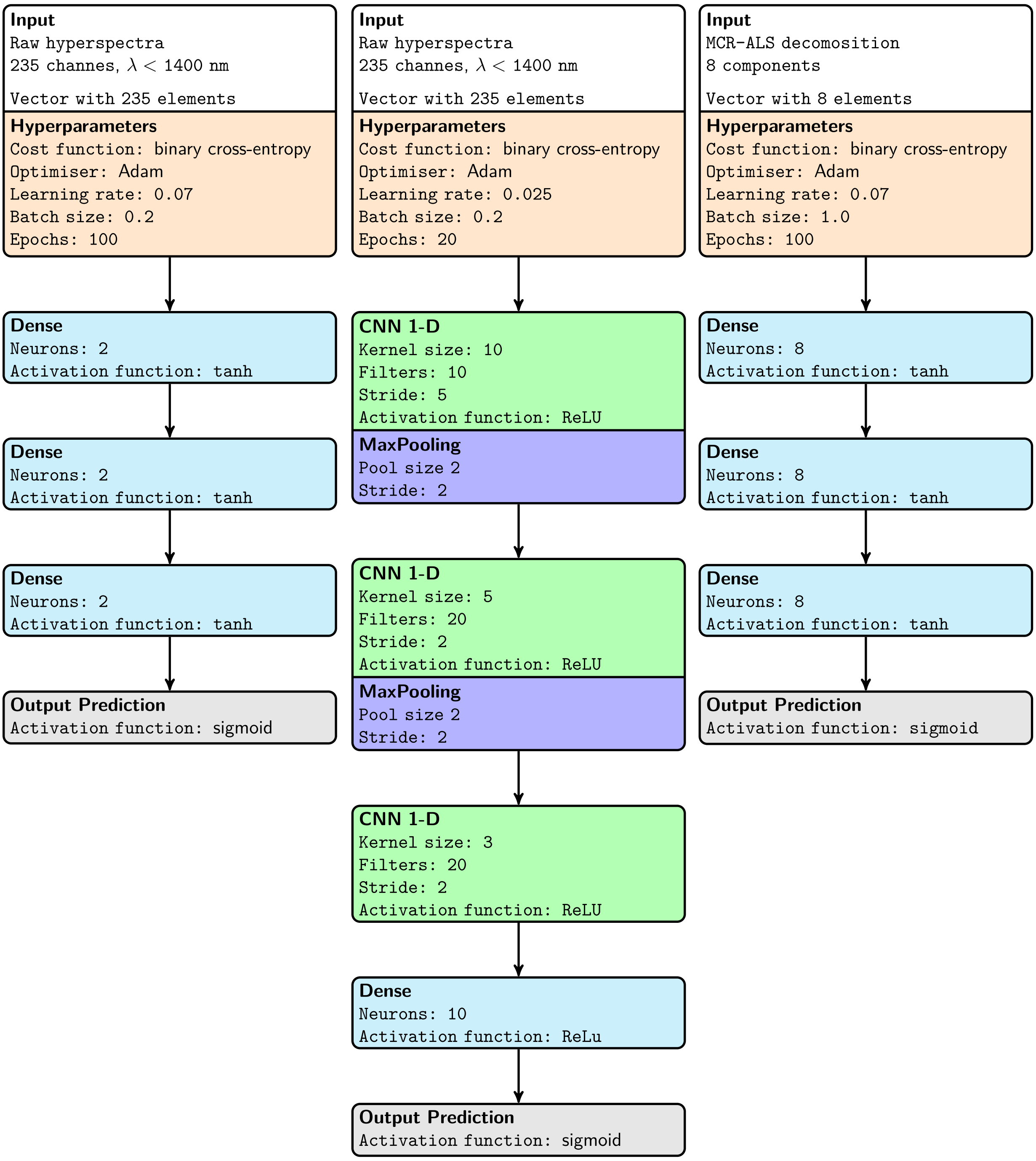

**Figure S4: Specifications of final models.** Model specifications for (left) the MLP, (middle) the CNN, and (right) the MLP on the MCR-ALS decomposition.

#### Sand-piles

This method can be formalized into a matrix problem as follows.

All the pixels has a predicted probability for being tumorous $p_i$. Those with probability $t_i = \mathbb{P}(p_i) \geq 0.5$ are considered to be tumorous. These are collected into a vector

$$\mathsf{t} = (t_1, t_2, \ldots, t_n), \tag{S1}$$

where $n$ is the number of pixels classified as tumorous. Only directly adjacent pixels are considered as neighbours, which can be summarized in a connectivity matrix

$$\mathsf{C} = \begin{pmatrix} 1 & w_{12} & \cdots & w_{1n} \\ w_{21} & 1 & \cdots & w_{2n} \\ \vdots & \vdots & \ddots & \vdots \\ w_{n1} & w_{21} & \cdots & 1 \end{pmatrix}, \tag{S2}$$

where

$$w_{ij} = \begin{cases} 1 & \text{if } j \in \text{neigh}(i), \\ 0 & \text{otherwise.} \end{cases} \tag{S3}$$

We want to construct a sand-pile

$$\mathsf{s} = (s_1, s_2, \ldots, s_n), \tag{S4}$$

where each element represents the height of the sand-pile on the corresponding pixel. During each iteration in the sand-pile process, a fraction of sand $b$ is moved from each pixel onto its neighbours. This is formalized with the update matrix

$$\mathsf{U} = \mathsf{C} - 4b\mathbb{1} = \begin{pmatrix} 1-4b & w_{12} & \cdots & w_{1n} \\ w_{21} & 1-4b & \cdots & w_{2n} \\ \vdots & \vdots & \ddots & \vdots \\ w_{n1} & w_{21} & \cdots & 1-4b \end{pmatrix}. \tag{S5}$$

No matter if a tumorous pixel has four or less tumorous neighbours, $4b$ is removed, i.e. the healthy pixels act as sinks. During one iteration of the sand-pile process, a unit of sand proportional to $t_i$ is also placed on each pixel $i$. One iteration can be summarized as

$$\mathsf{s}' = \mathsf{U}\mathsf{s} + \mathsf{t}, \tag{S6}$$

where $\mathsf{s}'$ is the updated sand-pile. This can equivalently be expressed with an augmented update matrix and the sand-pile vector augmented with "1":

$$\mathsf{M} = \left(\begin{array}{c|c} 1 & 0 \cdots 0 \\ \mathbf{t} & \mathsf{U} \end{array}\right) = \begin{pmatrix} 1 & 0 & 0 & \cdots & 0 \\ t_1 & 1-4b & w_{12} & \cdots & w_{1n} \\ t_2 & w_{21} & 1-4b & \cdots & w_{2n} \\ \vdots & \vdots & \vdots & \ddots & \vdots \\ t_n & w_{n1} & w_{21} & \cdots & 1-4b \end{pmatrix} \tag{S7}$$

and

$$\mathbf{v} = \begin{pmatrix} 1 \\ \mathbf{s} \end{pmatrix}, \tag{S8}$$

i.e.

$$\mathbf{v}' = \mathsf{M}\mathbf{v}. \tag{S9}$$

The 1 in the augmented vector $\mathbf{v}$ acts as a source together with the augmented column in $\mathsf{M}$. The zeroes in the top row are needed to not make the inflow of sand dependent on the amount of sand existing.

By repeatedly multiplying $\mathsf{M}$ with $\mathbf{v}$ a large number $N$ times, a stable state will be reached. We initialize the process without a pre-existing sand-pile

$$\begin{aligned} \mathbf{v}^{(0)} &= (1, 0, \ldots, 0)^T \\ \mathbf{v}^{(1)} &= \mathsf{M}\mathbf{v}^{(0)} \\ \mathbf{v}^{(2)} &= \mathsf{M}\mathbf{v}^{(1)} = \mathsf{M}(\mathsf{M}\mathbf{v}^{(0)}) = \mathsf{M}^2\mathbf{v}^{(0)} \\ &\vdots \\ \mathbf{v}^{(N)} &= \mathsf{M}\mathbf{v}^{(N-1)} = (\underbrace{\mathsf{M}\ldots\mathsf{M}}_{N \text{ times}})\mathbf{v}^{(0)} = \mathsf{M}^N\mathbf{v}^{(0)}. \end{aligned} \tag{S10}$$

Now, the problem has been reduced to finding $\mathsf{M}^N$ for $N \to \infty$. A straight-forward strategy would be to diagonalize $\mathsf{M}$ with $\mathsf{M} = \mathsf{PDP}^{-1}$ and find $\mathsf{M}^N$ by

$$\mathsf{M}^N = (\mathsf{PDP}^{-1})^N = \overbrace{(\mathsf{PD}\underbrace{\mathsf{P}^{-1})(\mathsf{P}}_{\mathbb{1}}\mathsf{DP}^{-1})\ldots(\mathsf{PDP}^{-1})}^{N \text{ times}} = \mathsf{PD}^N\mathsf{P}^{-1}, \tag{S11}$$

but this is computationally heavy. A more practical way which requires just a few computations is stepwise square $\mathsf{M}$, i.e.

$$\begin{aligned} \mathsf{M}^2 &= \mathsf{MM} \\ \mathsf{M}^4 &= \mathsf{M}^2\mathsf{M}^2 \\ \mathsf{M}^8 &= \mathsf{M}^4\mathsf{M}^4 \\ &\vdots \end{aligned} \tag{S12}$$

It does not require many iterations to obtain a large enough $N$. We iterate 10 times, yielding $N = 1024$.

#### SI Note III: MCR-ALS for all tumors

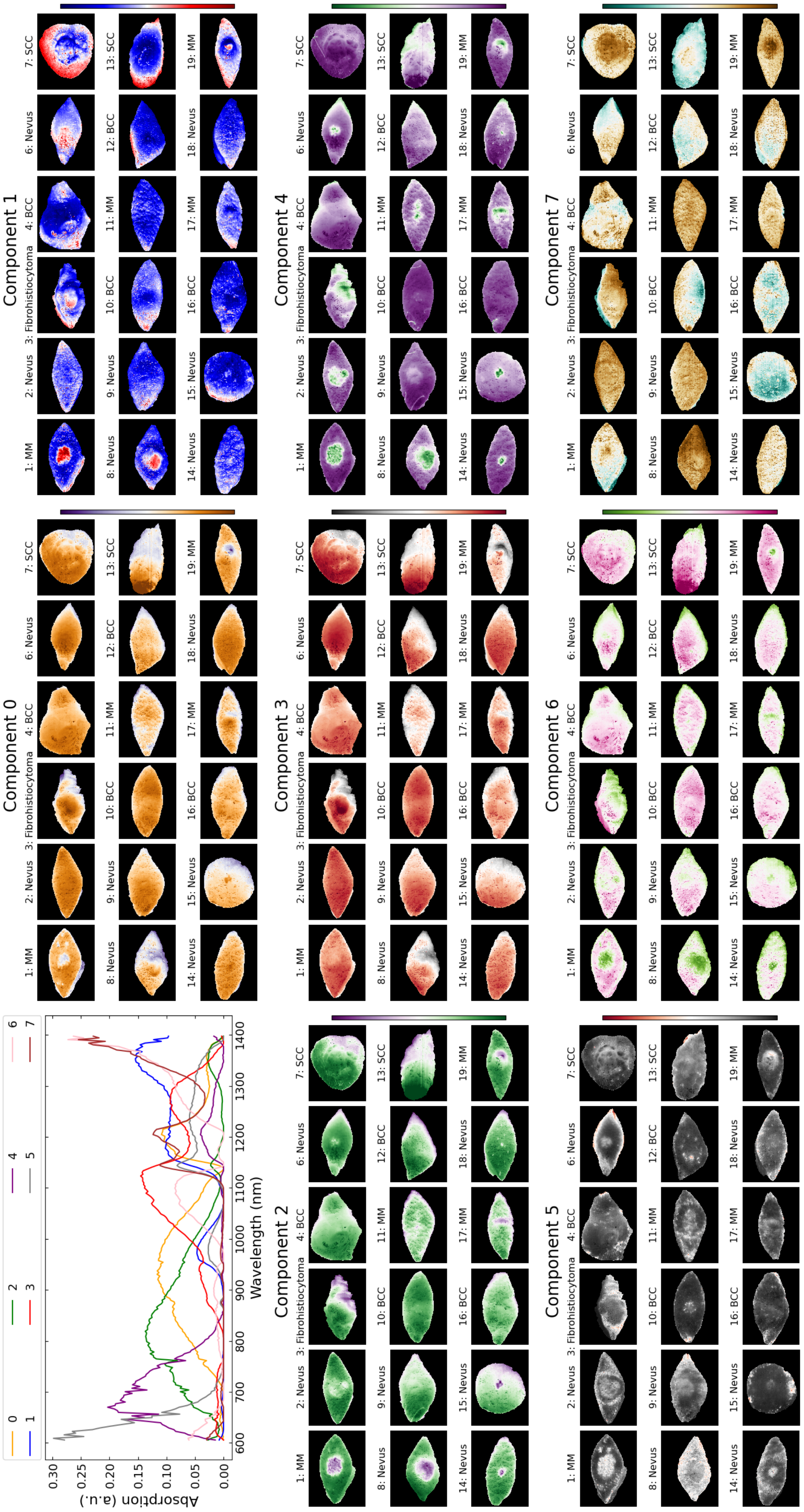

**Figure S5: MCR-ALS decomposition for all tumors.** The top-left panel shows the 8 spectral components, while the remaining 8 panels show the contribution of each component for all the tumor samples.

#### SI Note IV: Prediction maps for all tumors

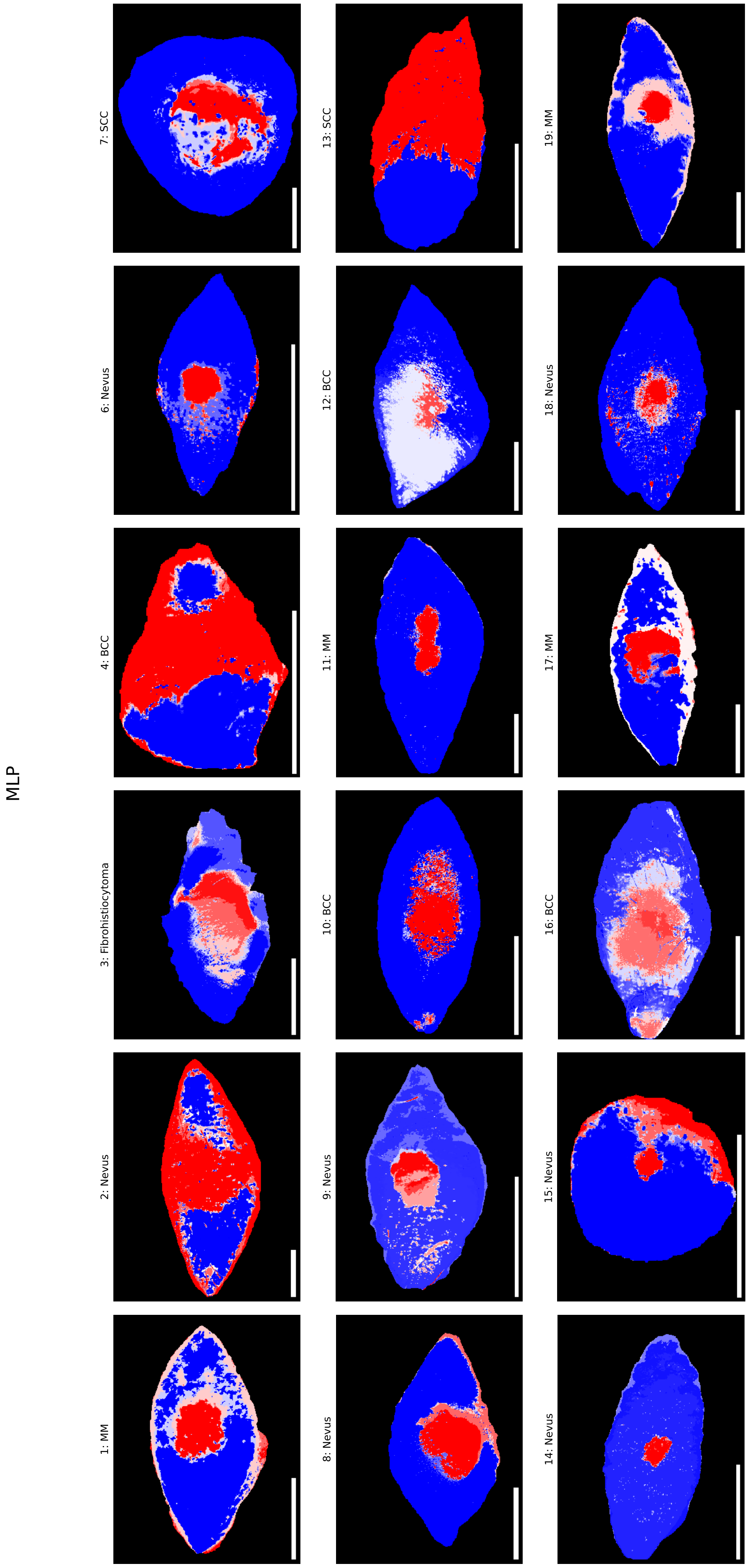

**Figure S6: Prediction map for the MLP.**

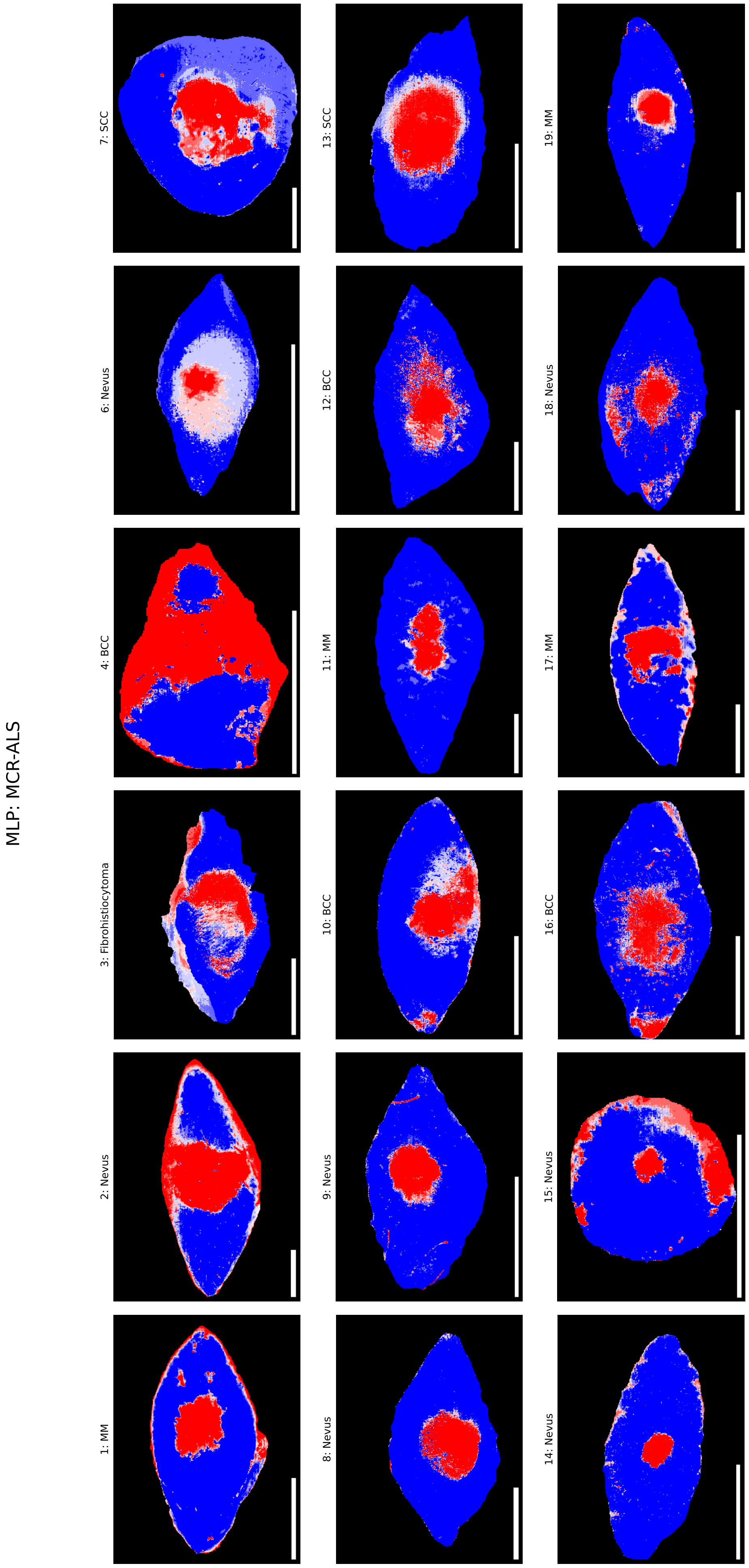

**Figure S7: Prediction map for the MLP: MCR-ALS.**

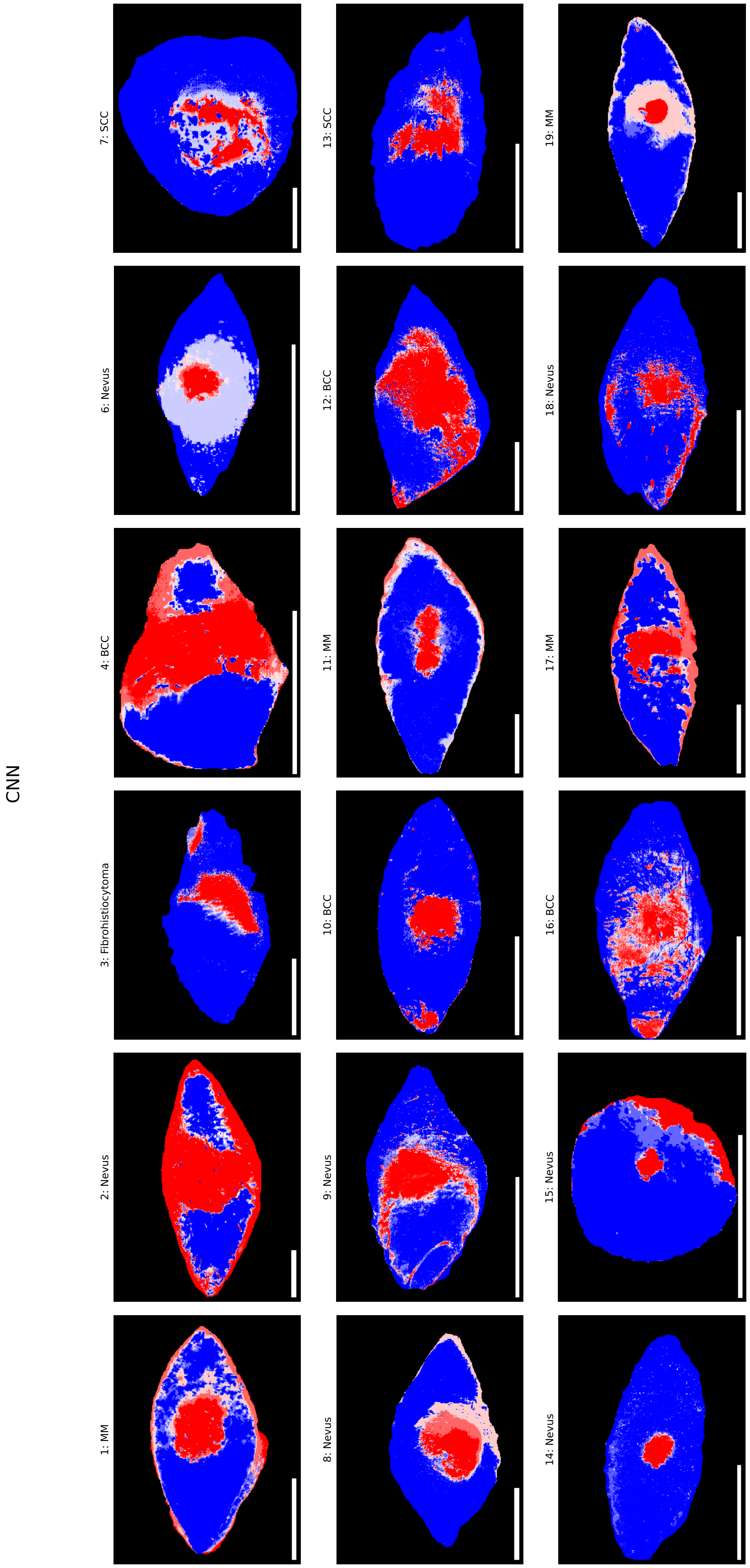

**Figure S8: Prediction map for the CNN.**

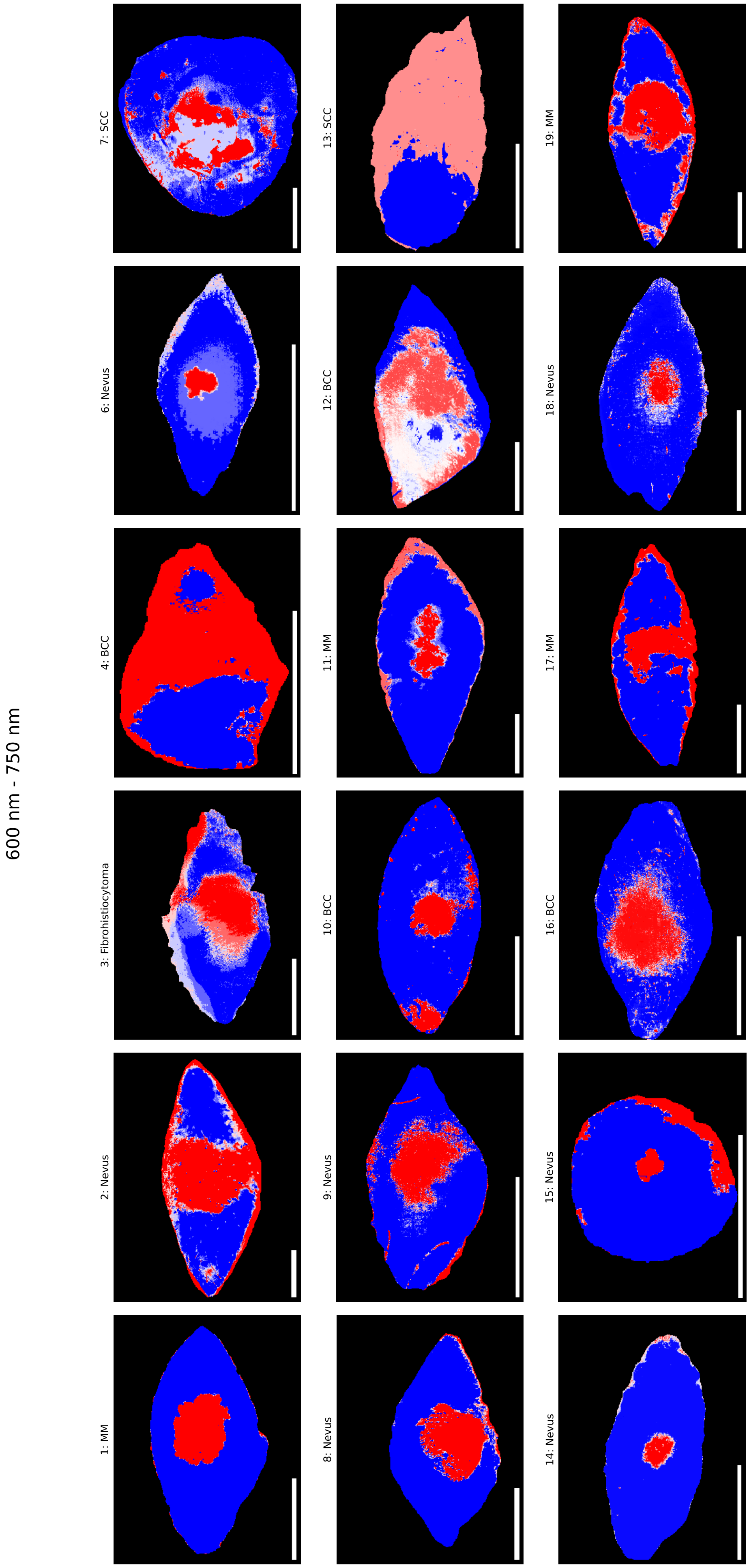

**Figure S9: Prediction map for the MLP in the spectral range 600 nm - 750 nm.**

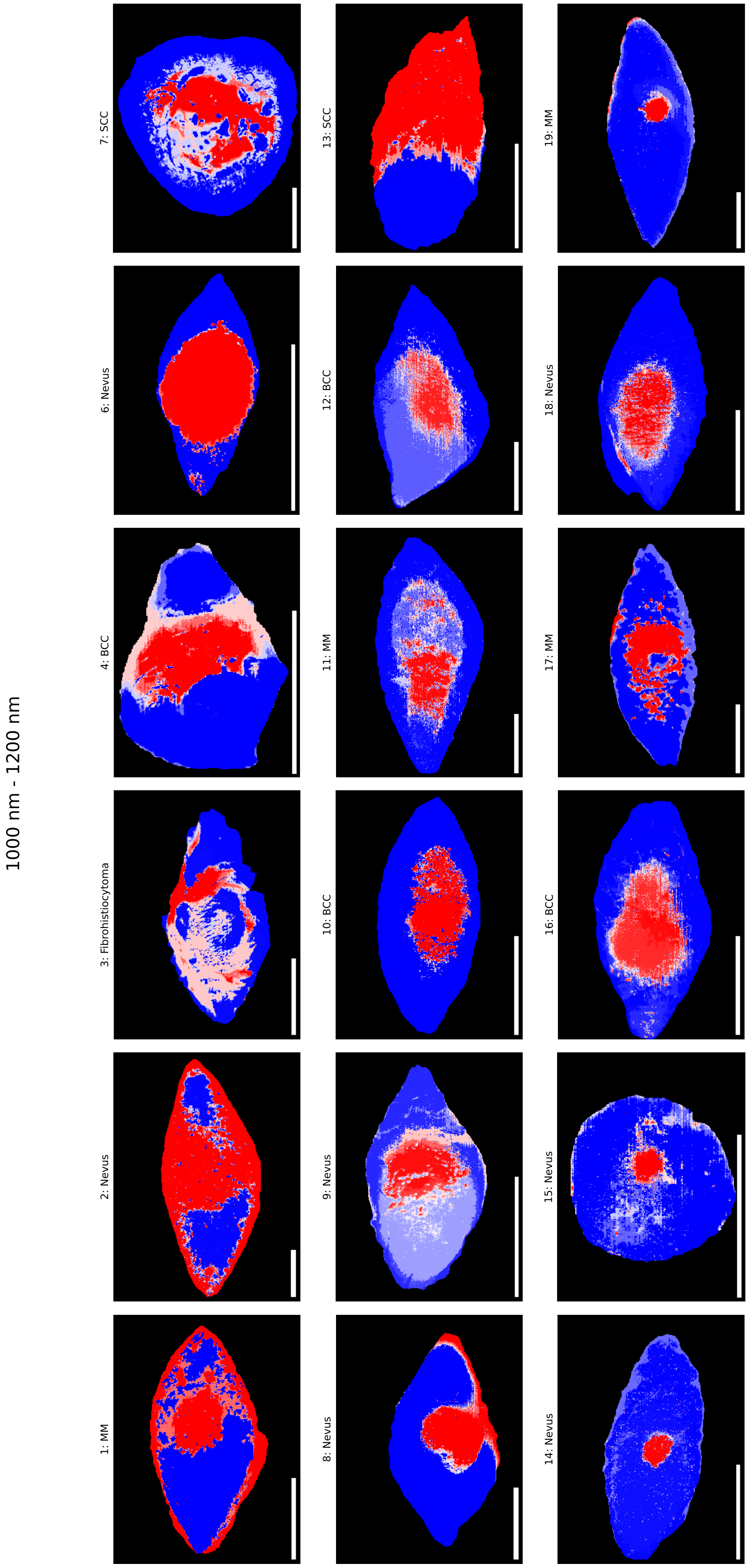

**Figure S10: Prediction map for the MLP in the spectral range 1000 nm - 1200 nm.**

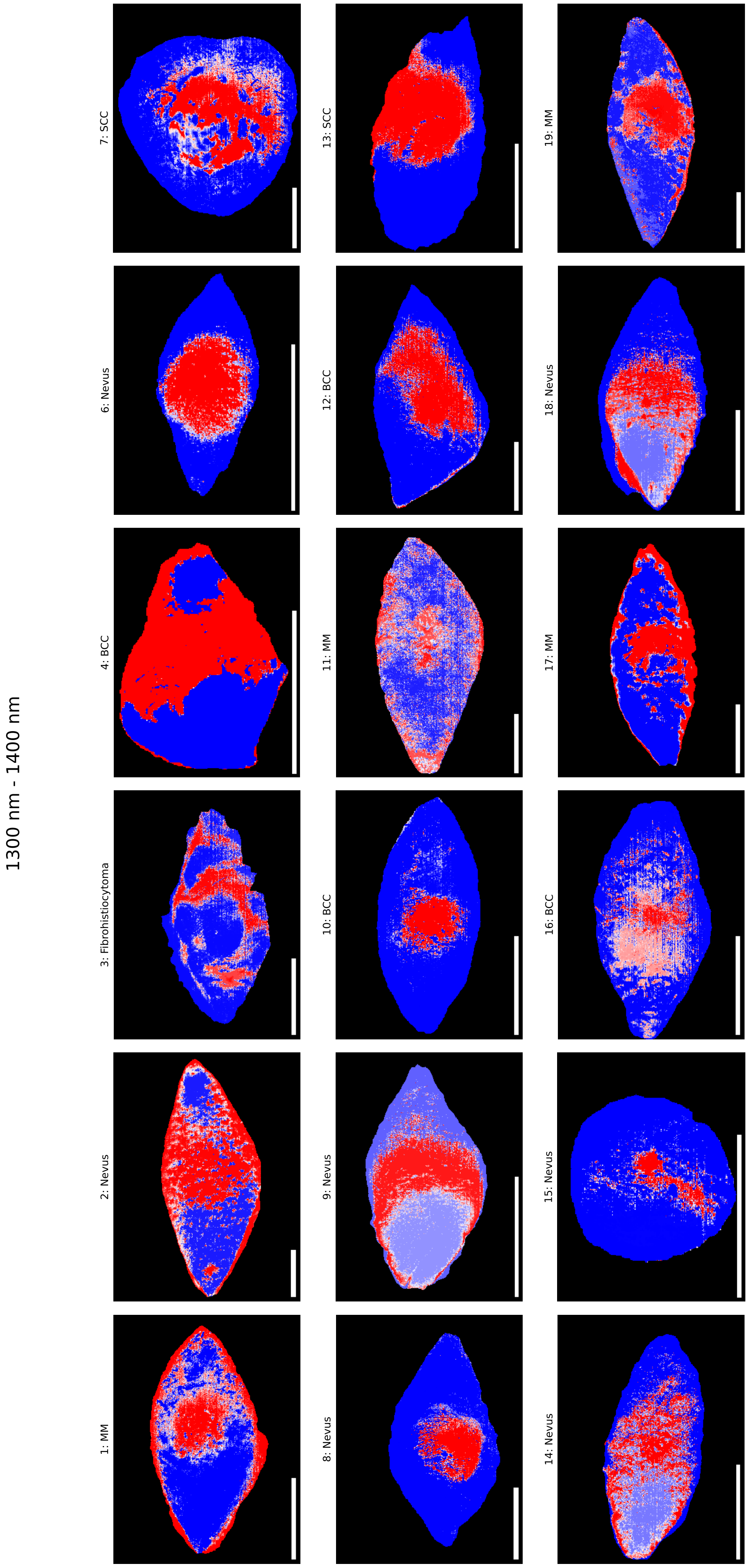

**Figure S11: Prediction map for the MLP in the spectral range 1300 nm - 1400 nm.**

### SI Note V: Model predicted tumor widths

**Table S4: Model widths.** Measured histology width and predicted tumor widths in mm for each of the 6 presented models for every tumor.

| Lesion no. | Histology | MLP | CNN | MLP: MCR-ALS | MLP: 600-750 nm | MLP: 1000-1200 nm | MLP: 1300-1400 nm |
|---|---|---|---|---|---|---|---|
| 1 | 6.89 | 6.90 | 7.20 | 5.95 | 7.10 | 7.75 | 7.10 |
| 2 | 15.7 | 15.8 | 15.8 | 15.7 | 15.9 | 13.0 | 13.1 |
| 3 | 6.78 | 6.90 | 6.45 | 6.95 | 6.25 | 5.80 | 6.65 |
| 4 | 7.83 | 5.65 | 3.35 | 3.30 | - | 4.60 | 3.45 |
| 6 | 2.39 | 2.36 | 2.67 | 2.77 | 1.96 | 5.53 | 4.33 |
| 7 | 13 | 16.0 | 13.4 | 16.5 | 14.2 | 19.8 | 19.9 |
| 8 | 9.45 | 9.25 | 9.25 | 8.25 | 9.15 | 8.65 | 8.65 |
| 9 | 4.89 | 3.77 | 6.13 | 4.17 | 5.12 | 6.08 | 5.17 |
| 10 | 3.45 | 5.26 | 5.01 | 5.01 | 4.56 | 5.61 | 5.11 |
| 11 | 8.58 | 4.40 | 4.35 | 6.00 | 5.70 | 9.45 | 2.55 |
| 12 | 6.76 | 4.55 | 11.6 | 7.61 | 12.1 | 7.40 | 9.71 |
| 13 | 4.97 | 7.70 | 6.49 | 6.69 | 8.15 | 7.85 | 4.68 |
| 14 | 2.86 | 2.60 | 2.91 | 2.96 | 2.91 | 2.86 | 6.28 |
| 15 | 1.6 | 1.75 | 1.70 | 1.85 | 1.75 | 2.15 | 2.30 |
| 16 | 6.61 | 6.25 | 7.10 | 5.60 | 5.75 | 6.10 | 6.60 |
| 17 | 8.31 | 8.35 | 8.20 | 8.35 | 8.30 | 8.35 | 7.40 |
| 18 | 4.84 | 4.05 | 4.60 | 4.35 | 3.10 | 5.05 | 6.85 |
| 19 | 8.54 | 9.90 | 9.80 | 7.15 | 10.7 | 4.60 | 10.3 |

#### SI Note VI: Saliency all tumors

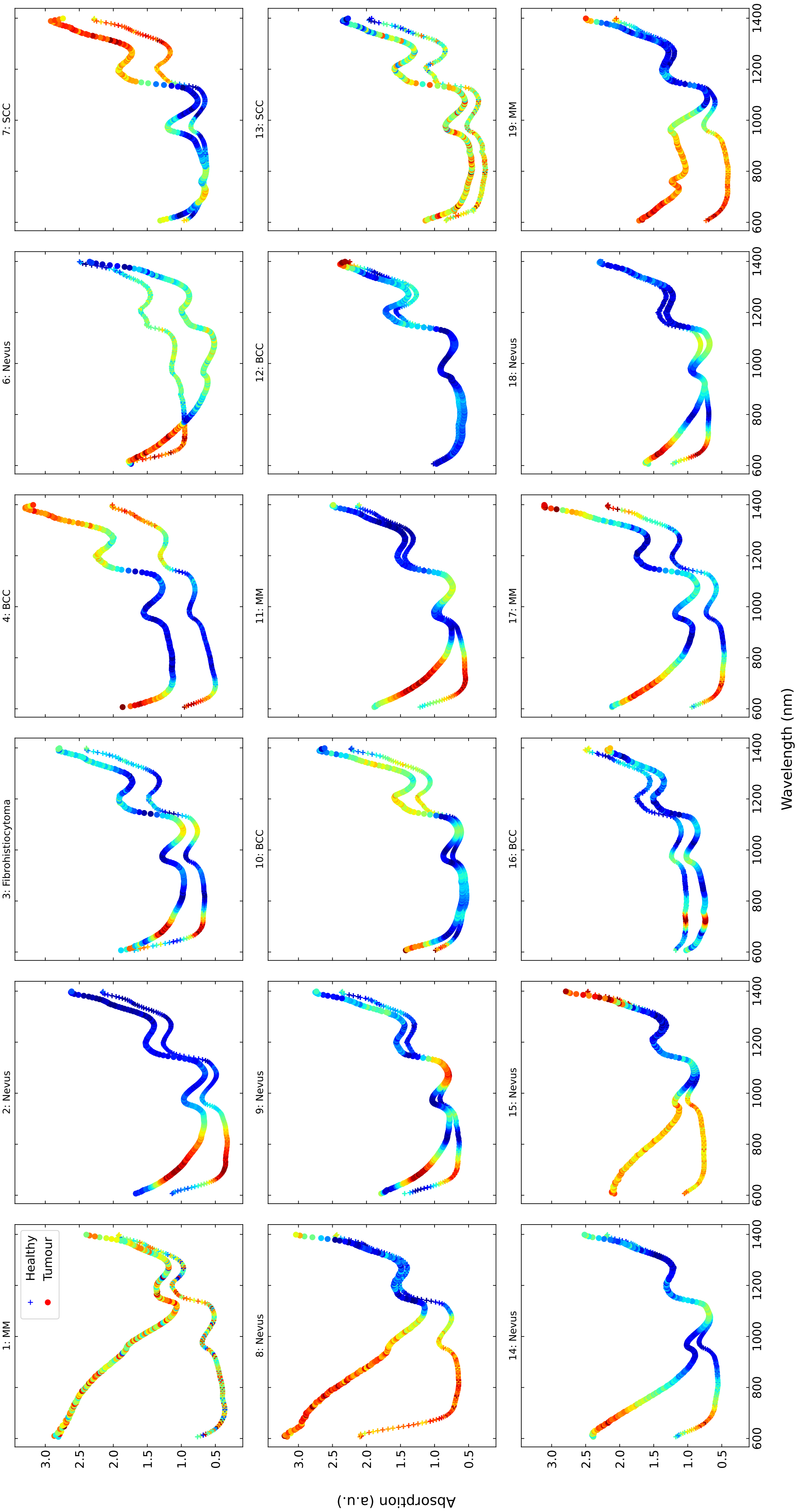

**Figure S12: Saliency for all the samples.**

### References

[1] Kingma, Diederik P., & Jimmy Ba. Adam: A method for stochastic optimization. *arXiv preprint* arXiv:1412.6980 (2014)
